# Food insecurity, adolescent suicidal thoughts and behaviours, and country-level context: a multi-country cross-sectional analysis

**DOI:** 10.1101/2023.06.15.23291417

**Authors:** Thomas Steare, Gemma Lewis, Sara Evans-Lacko, Alexandra Pitman, Kelly Rose-Clarke, Praveetha Patalay

## Abstract

**Background:** Preventing adolescent suicide is a global priority. Inequalities in adolescent suicide and attempt rates are reported across countries, including a greater risk in adolescents experiencing food insecurity. Little is known about the extent to which country-level contextual factors moderate the magnitude of socio-economic inequalities in suicidal thoughts and behaviours. We aimed to examine the cross-country variability and national moderators of the association between food insecurity and suicidal thoughts and behaviours in school-attending adolescents.

**Methods:** We analysed data on 309,340 school-attending adolescents from 83 countries that participated in the Global School-based Student Health Survey between 2003 and 2018. We used Poisson regression to identify whether suicidal thoughts and behaviours were more prevalent in adolescents experiencing food insecurity compared to food-secure adolescents. Meta-regression and mixed-effects regression were used to determine whether country-level indicators moderated the magnitude of inequality.

**Findings:** Suicidal ideation, suicide planning and suicide attempts were more prevalent in food-insecure adolescents compared to food-secure adolescents in 72%, 78%, and 90% of countries respectively; however, the magnitude of these associations varied between countries. We observed wider inequalities in countries with greater levels of national wealth and universal health coverage and lower prevalence of adolescent food insecurity. Economic inequality had no moderating role.

**Interpretation:** Food insecurity could contribute to the development of adolescent suicidal thoughts and behaviours, and this association is likely to be moderated by country-level context. Food insecurity may be a modifiable target to help prevent adolescent suicide, especially in countries where food insecurity is less common.

**Funding:** Wellcome Trust.

## Implications and Contributions

Suicidal ideation, planning, and attempts were more prevalent in food-insecure adolescents in 78% (65 of 83 countries), 72% (58 of 81 countries), and 90% (61 of 68 countries) of countries, respectively. The magnitude of inequality varied across countries, and in some countries, there was no evidence of disparities based on food insecurity. Inequalities were greater in countries where food insecurity among adolescents was uncommon, and in countries with higher levels of national wealth and universal health coverage. Our study highlights that inequalities in suicidal thoughts and behaviours according to food insecurity status are not universal and to some extent dependent on country-level context. Accordingly, policies and actions to tackle socio-economic inequalities in suicidal thoughts and behaviours and to ultimately prevent suicides need to consider the wider societal and policy context.

Suicide is a leading cause of death in adolescents.^1^ Its prevention is a global priority and a key target within the United Nation’s Sustainable Development Goal to promote health and well-being. ^2^ Over the past 20 years the decline in the rate of adolescent suicide has been marginal and in some countries rates have risen.^3^ Suicide risk is associated with an individual’s socio-economic position and higher in people experiencing forms of adversity, including food insecurity.^4,5^

Food insecurity, which refers to poor access to sufficient and nutritious food, affects more than two billion people worldwide.^6^ It is often a consequence of financial poverty, but non-financial factors like carer ill-health, political unrest, conflict, and agricultural performance also contribute. Food insecurity is associated with future adolescent mental health problems and suicidal ideation, independent of socio-demographic and parental confounders.^7,8^ Food insecurity therefore represents a modifiable and preventable stressor with far-reaching impacts on adolescent health.

Food insecurity may impact key cognitive-affective states that feature in models of suicidal thoughts and behaviour. One key psychological construct, thwarted belongingness, is characterised by unmet psychological needs for social connection and an absence of reciprocally-caring relationships.^9^ Adolescents experiencing food insecurity often report stigma and social isolation.^10^ Another relevant psychological construct is perceived burdensomeness, which refers to an individuals’ belief that they are a burden to others, especially family members, commonly co-occurring with distress and self-hate.^9^ Adolescents from food-insecure households report feeling shame, sadness, and family strain.^11^ If these experiences are perceived to be stable and unchanging and co-occur with a reduced fear of death, this can lead to active suicidal plans and behaviours.^9^

Social comparisons can negatively affect self-esteem and self-worth, leading to increased stress and mental health problems, and are likely to be exacerbated in contexts with large socio-economic inequalities.^12^ The impact of food insecurity on suicidal thoughts and behaviours may therefore depend on the level of economic inequality within a country, and how widespread food insecurity is.^5^ Socio-economic inequalities in adolescent psychological distress have been observed in European and North American countries with greater levels of income inequality,^13,14^ but it is unclear whether this extends to different mental health outcomes, countries, or social determinants.

The availability and accessibility of health services may also moderate the association between food insecurity and suicide thoughts and behaviours. Lower socio-economic position is associated with reduced access to primary mental healthcare, however inequalities in access are typically reduced in countries with universal healthcare which may reduce health inequalities.^15^

To date, research has primarily focused on the moderating role of national wealth on the association between food insecurity and suicide thoughts and behaviours. Results have been inconclusive, but two studies suggest that the association between food insecurity and suicide attempts may be weakest in low-income countries.^16,17^ Conclusions are limited due to the use of categorical World Bank income classifications, rather than continuous measures of national wealth. Additionally, several studies have adjusted for variables on the causal pathway meaning the magnitude of inequalities in suicide thoughts and behaviours has likely been underestimated.^5,16^

The Global School-based Student Health Survey (GSHS) is an ongoing cross-sectional school-based survey of adolescents conducted since 2003 across primarily low- and middle-income countries. Using GSHS data we examined cross-country variation in the magnitude of inequalities in adolescent suicidal thoughts and behaviours according to levels of food insecurity. We tested whether the magnitude of inequalities in suicidal thoughts and behaviours vary according to five country-level factors: the prevalence of food insecurity in adolescents, national wealth, income inequality, universal healthcare and the proportion of national health spending that is out-of-pocket.

## Methods

We pre-registered our protocol on the Open Science Framework (https://osf.io/kh4wx).

### Study design

We analysed cross-sectional data from the GSHS, available from 2003 to 2018. In almost all countries a two-stage stratified sampling design was used, with schools first selected with probability proportional to enrolment size.^18^ At the second stage, classes within selected schools were randomly chosen and all students within those classes were eligible to complete the survey during a regular class period. A census-based sampling approach was used in seven countries with a small population.

In each participating country, the study was approved by relevant national administrative bodies or ethics committees prior to data collection. Informed consent was secured before completion of the survey. The publicly available datasets featured anonymised data and no individually identifying information was collected. We used publicly available data and therefore no separate ethics approval was sought for our analyses.

At the time of analysis, data were available for 152 surveys across 104 countries. We included survey datasets with variables capturing food insecurity, suicidal ideation, planning and/or attempts, and where a national representative sampling framework was used. We used data from the most recent survey if there were multiple survey datasets within a country.

### Participants

We used complete case analysis. Participant data were analysed if data were available for food insecurity status, age, sex, and the specific suicidal behaviour. We did not impute missing data as missing data levels were low (<5%) and there were few variables that could be used to predict missing values in the imputation model, limiting potential efficiency gains or bias reduction.^19^

### Outcomes

The GSHS measures suicidal ideation, suicide planning and suicide attempts. Suicidal ideation and suicide planning were measured with two single items: “During the past 12 months, did you seriously consider attempting suicide?”, and “During the past 12 months, did you make a plan about how you would attempt suicide?” respectively. Responses for each item were either “yes” or “no.” Suicide attempts were measured with a single item: “During the past 12 months, how many times did you actually attempt suicide?”. We dichotomised responses: where adolescents reported one or more attempts, they were classed as “yes”; adolescents who reported no attempts were classed as “no”.

### Exposure

Food insecurity was assessed with a single item: “During the past 30 days, how often did you go hungry because there was not enough food in your home?”, with five response options: “never”, “rarely”, “sometimes”, “most of the time” & “always”.

We transformed the food insecurity measure using ridit scores. Ridit score transformation considers the population distribution across response categories (Supplementary Material, p2). Response categories with fewer respondents are indicative of a more extreme social position. In regression analyses of binary outcomes, ridit-transformed variables estimate the Relative Index of Inequality (RII), which represents the relative ratio of a binary outcome between those with the highest and lowest socio-economic position in a population.^20^

We also modelled food insecurity as a binary variable, in line with previous analyses.^21^ We followed the WHO’s dichotomisation in the GSHS Public Use Codebook (available for each country at https://extranet.who.int/ncdsmicrodata/index.php/catalog/GSHS) and classified adolescents as food-insecure if they reported going hungry “most of the time” or “always”.

We report the results of the RII analyses in the main text. We report the results of the binary exposure in the supplementary material (except for the moderation analysis of national prevalence of food insecurity which is presented in the main text).

### Confounders

We adjusted for age and sex (self-reported as male or female). To ensure cross-survey measurement comparability we used comparable age bandings: “13 years old or younger”, “14 years old”, “15 years old”, and “16 years old or older”. Other potential confounders including indicators of socio-economic background such as family income were not available in the GSHS and therefore were not controlled for.

### Country-level indicators

We estimated the weighted national prevalence of food insecurity among school-attending adolescents based on adolescents’ responses on the food insecurity item. This estimate refers to the percentage of the country sample who reported being food-insecure “most of the time” or “always” in the past 30 days.

We accessed data on further country-level indicators from the World Bank Data Catalog (https://datacatalog.worldbank.org/home). We drew indicator data from the corresponding survey year for each country. If country-level data were not available for the specific survey year, we used data closest to the relevant year going backwards in time.

We used countries’ Gross Domestic Product (GDP) per capita expressed in 2021 international dollars as an estimate of economic prosperity. GDP refers to the sum gross value added by a country’s residents, with purchase power parity applied to control for cross-country price differences in goods. In a deviation from our protocol, we log-transformed GDP per capita. The impact of an increase in GDP per capita on a nation’s development is likely to diminish as GDP per capita increases, and thus has a non-linear impact.^22^

We used the Gini index as a measure of economic inequality within each country. The Gini index measures the equality of a country’s income distribution, with a higher value indicating greater income inequality.

We used measures we considered to be suitable proxies for the availability of accessible free healthcare. We used the WHO Universal Health Coverage Index. The Index is based on sixteen different indicators from four domains of healthcare. It is important to note that no indicators specifically relate to mental health services, and although overall healthcare access might relate to availability of mental health services, given the limited mental healthcare in many low- and middle-income countries this might not be the case and general healthcare services might be the main provider of mental healthcare. We also accessed data on the level of out-of-pocket healthcare expenditure, excluding taxes and insurance premiums, as reported in the WHO Global Health Expenditure database. Data represent the percentage of a country’s health expenditure that is directly paid for by households.

### Analysis

We ran Poisson regression models for each country, exposure (ridit-transformed and binary) and outcome (suicidal ideation, planning, and attempt). We adjusted for sex and age. We accounted for country specific primary sampling units (classrooms), strata (school) and sample weights.

We pooled results from country-level Poisson regression models into a random-effects meta-analysis to identify the cross-country variability in RIIs and prevalence ratios. We used restricted maximum likelihood estimation to identify the level of total variation in RIIs and prevalence ratios between countries due to heterogeneity rather than chance (*I*^2^ statistic). We ran random-effects meta-regression analyses testing for the moderating role of each country-level indicator.

As a complementary analysis we ran mixed-effects Poisson regression models for the pooled sample to test for interactions between food insecurity and each country-level indicator using R package *lme4*. There was no option to include sampling weights, leading to a deviation from our protocol. In all models, food insecurity, age, and sex added as fixed effects, countries as random effects, and food insecurity included as a random slope. To test for interactions between food insecurity and country-level indicators, separate models were run for each individual country-level indicator (with a main effect for country-level indicator and an interaction with food insecurity).

For all meta-regression and mixed-effects analyses, moderators were adjusted for national wealth. We additionally adjusted for Gini coefficients in models where the prevalence of food insecurity among school-attending adolescents was the country-level indicator of interest. For the mixed-effects analyses, we standardised country-level indicator data to reduce potential multi-collinearity in the models (except log-transformed GDP per capita which was mean-centred to help model convergence).

Ridit scores are calculated according to the distribution of food insecurity, meaning any association between the RII and the prevalence of food insecurity among school-attending adolescents would be partially tautological. We therefore did not look at the moderating role of national food insecurity prevalence on the RIIs.

### Sensitivity analyses

We re-ran all analyses for suicidal ideation and suicide planning using data from the 68 countries where all outcomes were measured, to assess whether potential differences in results across outcomes may be influenced by the number of, or specific countries included in each analysis.

We ran additional Poisson regression models at country-level for each outcome and exposure using data from surveys previously excluded as there were more recent datasets used in the main analysis for that specific country. We compared results of the analyses of the additional and existing survey datasets for each country to identify whether the magnitude of inequalities was consistent over time within countries.

### Role of the funding source

The funder had no role in the study design, the interpretation of data, the writing of this manuscript, or the decision to submit for publication.

## Results

We analysed data on an eligible sample of 315,505 school-attending adolescents with data on key measures from 83 countries and territories. Our samples representing suicidal ideation, suicide planning and suicide attempts consisted of 309,340, 303,135, and 277,181 adolescents, from 83, 81, and 68 countries respectively (Supplementary Figure 1 & 2). The proportion of participants excluded owing to missing data was 4·7% for the suicidal ideation sample, 4·7% for the suicide planning sample, and 3·3% for the suicide attempt sample.

Over a third of participants were from the Americas, including 23 countries from that region (Table 1). We included eight overseas territories; one in the Western Pacific Region (Cook Islands), whilst seven have not been classified by the WHO.

**Table 1.**
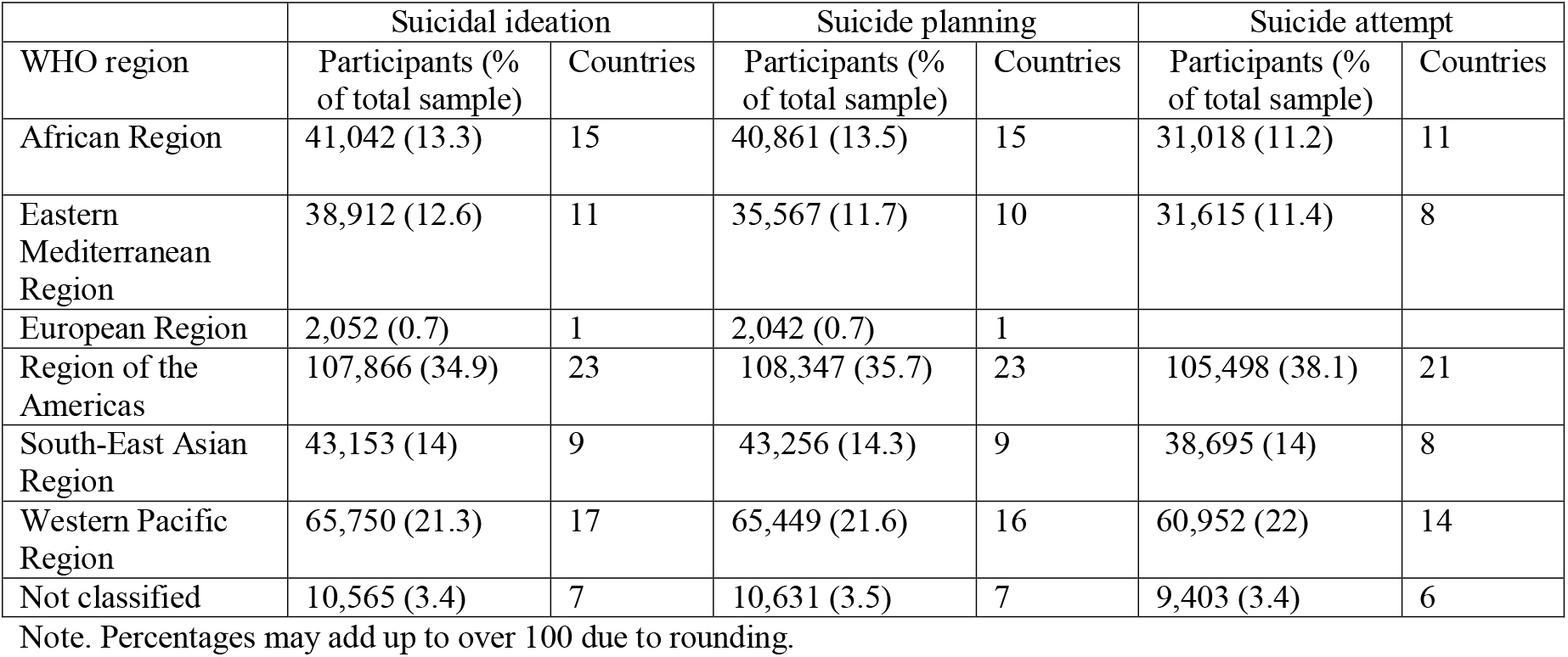
Participants according to WHO region.

| WHO region | Suicidal ideation |  | Suicide planning |  | Suicide attempt |  |
| --- | --- | --- | --- | --- | --- | --- |
|  | Participants (% of total sample) | Countries | Participants (% of total sample) | Countries | Participants (% of total sample) | Countries |
| African Region | 41,042 (13.3) | 15 | 40,861 (13.5) | 15 | 31,018 (11.2) | 11 |
| Eastern Mediterranean Region | 38,912 (12.6) | 11 | 35,567 (11.7) | 10 | 31,615 (11.4) | 8 |
| European Region | 2,052 (0.7) | 1 | 2,042 (0.7) | 1 |  |  |
| Region of the Americas | 107,866 (34.9) | 23 | 108,347 (35.7) | 23 | 105,498 (38.1) | 21 |
| South-East Asian Region | 43,153 (14) | 9 | 43,256 (14.3) | 9 | 38,695 (14) | 8 |
| Western Pacific Region | 65,750 (21.3) | 17 | 65,449 (21.6) | 16 | 60,952 (22) | 14 |
| Not classified | 10,565 (3.4) | 7 | 10,631 (3.5) | 7 | 9,403 (3.4) | 6 |

Note. Percentages may add up to over 100 due to rounding.

The prevalence of food insecurity and of suicidal thoughts and behaviours varied across the 83 countries and territories (Supplementary Table 1). Across the pooled sample, 6·4% of adolescents reported food insecurity in the past 30 days (Supplementary Table 2). Suicidal ideation, suicide planning and suicide attempts in the previous 12 months were reported by 12·6%, 11·7%, and 11·3% of participants respectively and were moderately correlated with each other (Supplementary Table 3 & 4).

### Inequalities in adolescent suicidal thoughts and behaviours according to food insecurity status

#### Suicidal ideation

The prevalence of suicidal ideation was lower in the most food-secure adolescents compared to the least food-secure adolescents in 65 of the 83 countries (Figure 1 & 2). For the pooled sample the RII was 0·51 (95% CI: 0·47 to 0·55), and there was evidence of variability in the magnitude of inequality across countries. The RII was greatest in Myanmar (RII=0·15, 95% CI: 0·09 to 0·25), with the prevalence of suicidal ideation 85% lower in the most food-secure adolescents compared to the least food-secure adolescents. The *I*^2^ statistic was 72·6%, indicating a substantial proportion of the total variability in RIIs was due to between-country heterogeneity.

#### Suicide planning

In 58 of the 81 countries represented for this outcome, the prevalence of suicide planning was lower in the most food-secure adolescents compared to the least food-secure adolescents (Figure 1 & 2). The RII was 0·55 (95% CI: 0·51 to 0·59) in the pooled sample, but varied across countries. The RII was greatest in North Macedonia (RII=0·25, 95% CI: 0·13 to 0·47). The *I*^2^ statistic was 64·1%, indicating a substantial proportion of the total variability in RIIs for suicide planning was due to between-country heterogeneity.

#### Suicide attempt

In 61 of the 68 countries represented for this outcome, the prevalence of attempts was lower in the most food-secure adolescents compared to the least food-secure adolescents (Figure 1 & 2). The RII was 0·45 (95% CI: 0·42 to 0·5) in the pooled sample. The RII varied across countries and was greatest in Morocco (RII=0·2, 95% CI: 0·16 to 0·25). The *I*^2^ statistic was 71·6% indicating a substantial proportion of the total variability in RIIs were due to between-country heterogeneity.

### Country-level indicators as moderators

The proportion of total variance of the outcome attributable to differences between countries was estimated to be 8·4% for suicidal ideation, 8·7% for suicide planning, and 9·6% for suicide attempts as reported by the intraclass correlation coefficient for the mixed-effects models with no fixed effects entered, and a random intercept only.

Most country-level indicators were only weakly correlated with one another (Supplementary Table 5). The UHC index had a moderate correlation with GDP per capita (r=0·61) and with the national prevalence of food insecurity (r=-0·53).

#### National prevalence of food insecurity among school-attending adolescents

The national prevalence of food insecurity among school-attending adolescents moderated the association between the binary measure of food insecurity and all three outcomes in the meta-regression (Supplementary Table 6 & 7) and mixed-effects analyses (Supplementary Table 8). Prevalence ratios were typically higher in countries where food insecurity was less prevalent (Figure 3). According to the meta-regression analyses, food insecurity explained approximately 55% of between-country variability for suicidal ideation and suicide planning, and 77·8% for suicide attempts. Findings were robust after controlling for GDP per capita and income inequality.

#### GDP per capita (log-transformed)

GDP per capita moderated the RII for suicidal ideation in the meta-regression (Table 3) and mixed-effects analyses (Supplementary Table 9). Inequality in suicidal ideation was greater in countries with higher levels of GDP per capita. GDP per capita did not moderate the RII for suicide planning or attempts. According to the meta-regression analyses, GDP per capita explained approximately 10·7% of between-country variability for suicidal ideation, 3·8% for suicide planning, and 4·7% for suicide attempts.

#### Country-level inequality

Income inequality did not moderate the RII for any outcome. Income inequality explained almost none of the between-country variance in the meta-regression analyses.

#### Universal Health Coverage index

The level of universal health coverage was found to moderate the RII for all three outcomes in both the meta-regression (Table 2) and mixed-effects analyses (Supplementary Table 9). The RII was lower in countries with a higher level of universal health service coverage. In the meta-regression analyses the UHC index explained between 14 and 18% of the between-country variability for all outcomes. Findings were robust after controlling for GDP per capita.

**Table 2.** Meta-regression analyses of country-level indicators associated with the magnitude of country-level Relative Index of Inequality.

|  | GDP per capita |  |  | Gini index |  |  | Universal Health Coverage index |  |  | Out-of-pocket health spending |  |  |
| --- | --- | --- | --- | --- | --- | --- | --- | --- | --- | --- | --- | --- |
|  | Countries | Change in RII per increase in log dollars in GDP per capita | Between-country variance explained (%) | Countries | Change in RII per 1-point increase in the Gini index | Between-country variance explained (%) | Countries | Change in RII per 1-point increase in the UHC index | Between-country variance explained (%) | Countries | Change in RII per 1 percentage point increase in out-of-pocket health spending | Between-country variance explained (%) |
| Suicidal ideation |  |  |  |  |  |  |  |  |  |  |  |  |
| Unadjusted | 76 | -0.1<br>(-0.278 to -0.0238) | 10.7 | 55 | -0.003207<br>(-0.016 to 0.00953) | 0 | 74 | -0.00919<br>(-0.0146 to -0.0038) | 17.9 | 74 | -0.00334<br>(-0.00747 to 0.000796) | 3 |
| Adjusted<sup>a</sup> |  |  |  | 55 | -0.00184<br>(-0.0136 to 0.00994) | 18.4 | 74 | -0.0129<br>(-0.0226 to -0.00314) | 16.5 | 74 | -0.00413<br>(-0.00816 to -0.000104) | 13.9 |
| Suicide planning |  |  |  |  |  |  |  |  |  |  |  |  |
| Unadjusted | 74 | -0.069<br>(-0.141 to 0.0003) | 3.8 | 54 | -0.00338<br>(-0.0144 to 0.00762) | 0 | 72 | -0.00712<br>(-0.0121 to -0.00218) | 14.1 | 72 | -0.00276<br>(-0.0065 to 0.00098) | 2.7 |
| Adjusted<sup>a</sup> |  |  |  | 54 | -0.0025<br>(-0.0129 to 0.00794) | 12.9 | 72 | -0.0122<br>(-0.021 to -0.00346) | 16.6 | 72 | -0.00321<br>(-0.00695 to 0.000518) | 6.7 |
| Suicide attempt |  |  |  |  |  |  |  |  |  |  |  |  |
| Unadjusted | 61 | -0.0805<br>(-0.173 to 0.0234) | 4.7 | 46 | -0.00187<br>(-0.018 to 0.0142) | 0 | 60 | -0.00974<br>(-0.0164 to -0.00306) | 16.7 | 60 | -0.00359<br>(-0.00833 to 0.00115) | 3.3 |
| Adjusted<sup>a</sup> |  |  |  | 46 | 0.00221<br>(-0.0128 to 0.0173) | 19.4 | 60 | -0.0156<br>(-0.0272 to -0.00412) | 17.9 | 60 | -0.00402<br>(-0.0087 to 0.000655) | 8.4 |

<sup>a</sup>adjusted for log GDP per capita (PPP). Coefficients and 95% confidence intervals are presented. A positive coefficient indicates that an increase in the country-level indicator is associated with a reduced magnitude of inequalities in suicidal thoughts and behaviours according to food insecurity. A negative coefficient indicates that an increase in the country-level indicator is associated with an increased magnitude of inequalities in suicidal thoughts and behaviours according to food insecurity.

#### Out-of-pocket health expenditure

The level of out-of-pocket health expenditure did not moderate the RII of any outcome in the unadjusted analyses. Out-of-pocket health expenditure explained a minimal amount of the between-country variability in the RII (between 3% and 8% across outcomes). When adjusting for GDP per capita, the RII for suicidal ideation was greater in countries with higher levels of out-of-pocket health expenditure.

### Binary measure of food insecurity

Results for analyses using a binary measure of food insecurity are presented in the supplementary material (Supplementary Tables 6-8; Supplementary Figure 3). Results were largely consistent with those of the RII analyses, but GDP per capita additionally moderated the association between the binary food insecurity measure and suicide planning and attempts.

### Sensitivity analyses

Results were very similar after limiting the analyses to countries where data on all outcomes were available, however we no longer found a moderating role of GDP on the RII of suicidal ideation in the mixed-effects analyses (Supplementary Tables 10-13). This suggests we may not have identified a moderating role of GDP on suicide planning or attempts due to fewer countries included (hence reduced power) in those outcome-specific analyses. Within-country comparisons across different survey years indicated that RIIs were predominantly consistent over time for all outcomes, as indicated by overlapping 95% confidence intervals in the within-country estimates (Supplementary Figures 4 & 5).

## Discussion

Adolescents experiencing food insecurity are more likely than food-secure adolescents to report suicidal thoughts, make a suicide plan or make a suicide attempt in most countries sampled (more than 70% for each outcome). The magnitude of inequality varied between countries, with large differences in the prevalence of suicide thoughts and behaviours between food-insecure and food-secure adolescents in some countries, and small or no detectable differences in others. Differences in the extent of inequality between food-secure and food-insecure adolescents may be due to differing social and economic context; with our analyses showing the potential importance of the prevalence of food insecurity, national wealth, and the level of universal healthcare within a country.

Food insecurity was more strongly associated with each outcome in countries where the prevalence of food insecurity was low, building on previous analyses of suicide attempts only.^5^ This suggests that the psychological and social effects and correlates of food insecurity may be intensified where food insecurity is rare and more indicative of a reduced standard of living and low social standing within a country. In communities where food insecurity is common, social norms for food-insecure adolescents may be referenced to other food-insecure adolescents, rather than peers with a higher standard of living.^23^ In these contexts, food insecurity may have a lesser impact on thwarted belongingness, and in turn on risk of suicidal thoughts and behaviours.

Our findings suggest that the magnitude of inequality in suicidal ideation according to food insecurity status is greater in countries with higher levels of GDP. We build upon previous analyses of the GSHS that reported inconclusive results, partially limited by their categorisation of income data.^5,16,17^ The moderating role of national wealth remains unclear, except through its role as an important antecedent of other country-level factors.^23^

Counterintuitively, inequality according to food insecurity status was greater in countries with greater coverage of universal health services. We expect that this is likely to be driven by co-occurring low levels of food insecurity, or other correlated country-level factors. We found no moderating role for national out-of-pocket health care expenditure. Out-of-pocket health expenditure and the UHC index were only weakly correlated despite our assumption that a greater proportion of health expenditure is out-of-pocket where there is less universal healthcare. This suggests that the measures may not be appropriate proxies for mental healthcare accessibility in low- and middle-income countries. Our findings are consistent with research showing that health inequalities are not lower in countries with substantial welfare policies and higher expenditure on health services. ^24^

It is hypothesised that in economically unequal environments, negative social comparisons, which affect one’s self-esteem and mental health, are more salient, therefore potentially exacerbating socio-economic inequalities in in suicidal thoughts and behaviours.^12^ Contrary to our hypotheses, we found no moderating role of national income inequality as represented by the Gini coefficient for any outcome. Previous studies in higher income countries have demonstrated moderation by income inequality for economic inequalities in adolescent mental health.^13,14^ This might not extend to low- and middle-income settings or to food insecurity, and our findings highlight the importance of not assuming findings from high-income countries extend to all settings.

Our study has various strengths. We used data from a wide range of countries that are underrepresented in mental health research despite most of the global adolescent population living in low- or middle-income countries. In comparison to previous analyses, we modelled food insecurity and national wealth as continuums, rather than arbitrary binary variables and thus our findings should have greater validity.

However, the following limitations need to be considered. First, we used cross-sectional data and were unable to control for potential unmeasured confounders such as parental health. Second, health literacy in adolescents varies according to socio-economic background,^25^ and the interpretation of measures may have differed between food-secure and food-insecure adolescents. Third, we found variability in the prevalence of suicidal thoughts and behaviours across countries. This variability is likely to be partially driven by cross-country differences in measurement interpretation and cultural understandings, societal stigma, and national laws regarding suicide leading to potential over- or under-reporting. Fourth, we analysed data on school-attending adolescents, which may have introduced collider bias. In many participating countries adolescent school enrolment is low, and we therefore excluded adolescents not attending school, whose risk of experiencing food insecurity and suicide thoughts or behaviours may be higher. Finally, potentially more suitable country-level indicators might have been considered, such as variables capturing social protection policies or national suicide prevention strategies. However, these indicators do not exist in the GSHS dataset or were limited to a small number of countries.

Evidence from our cross-country analyses highlight the relevance of food insecurity in the development of suicidal thoughts and behaviours, and therefore could be an important target for adolescent suicide prevention. Longitudinal cross-country research is needed to investigate the timing and developmental trajectory of this association to establish the causal relationship. Targeted suicide prevention strategies for food-insecure adolescents may be more effective in higher income countries where food insecurity is uncommon. Importantly, given the increased inequalities seen in high-income countries, and the higher prevalence of food insecurity seen in low- and middle-income countries, tackling adolescent food insecurity should be a global priority with likely wide-reaching economic, social and health benefits.

## Supporting information

STROBE checklist

## Data Availability

Data were accessed from, and are available through the WHO NCD Microdata Repository: https://extranet.who.int/ncdsmicrodata/index.php/catalog/GSHS

## Declarations of interest

TS is funded by a UCL-Wellcome Trust doctoral training fellowship in mental health science. AP is funded by the NIHR UCLH Biomedical Research Centre.

## Acknowledgements

This paper uses data from the Global School-Based Student Health Survey (GSHS). GSHS is supported by the World Health Organization and the US Centers for Disease Control and Prevention. This work was supported by the Wellcome Trust [218497/Z/19/Z]. The MRC Unit for Lifelong Health and Ageing at UCL is funded by the Medical Research Council: MC_UU_00019/3 Programme 2: Mental Ageing.

## Notes

### Competing Interest Statement

The authors have declared no competing interest.

### Funding Statement

Funding: Wellcome Trust. The funder had no role in the study design, the interpretation of data, the writing of this manuscript, or the decision to submit for publication. No authors received payment or services from a third party for any aspect of this work.

