## Supplementary material for "Food insecurity, adolescent suicidal thoughts and behaviours, and country-level context: a multi-country cross-sectional analysis"

### **Details of ridit scoring and the Relative Index of Inequality**

For each country, responses on the food insecurity measure were transformed using ridit scoring. For each item response, transformation involved adding the cumulative proportion of responses in all lower categories to one-half of the proportion of cases within the responses own corresponding category. We classified “always” to be the lowest response, and “never” as the highest response for the food insecurity measure. The ridit-transformations model food insecurity on a continuum rather than an arbitrary binary grouping, therefore, analyses with the ridit-transformed exposure should have greater statistical power and validity as an arbitrary cut-off is no longer used to define food insecurity. Ridit-transformations allow the estimation of the Relative Index of Inequality, which is commonly used to compare the magnitude of health inequalities across countries or populations.

### **Deviations from protocol**

1. We used log-transformed values of GDP per capita (Purchase Power Parity) as an indicator of national wealth, instead of raw values. For the mixed-effects analyses these values were then also mean-centred because of non-convergence warnings when using the log-transformed values as fixed effects in the models.

2. We ran mixed-effects analyses in R using the package *lme4* which has no option to include sampling weights. Mixed-effects analyses were subsequently run without sampling weights.

3. We reported the ICC for the null mixed-effects models only, as the ICC for mixed-effects Poisson models becomes difficult to meaningfully interpret once fixed effects are added.

### **Supplementary Figure 1. Sample flowchart of GSHS countries and participants.**

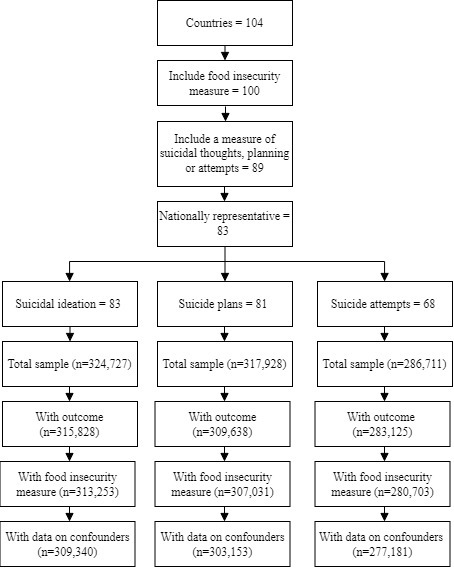

### **Supplementary Figure 2. Map of GSHS countries included in the analysis.**

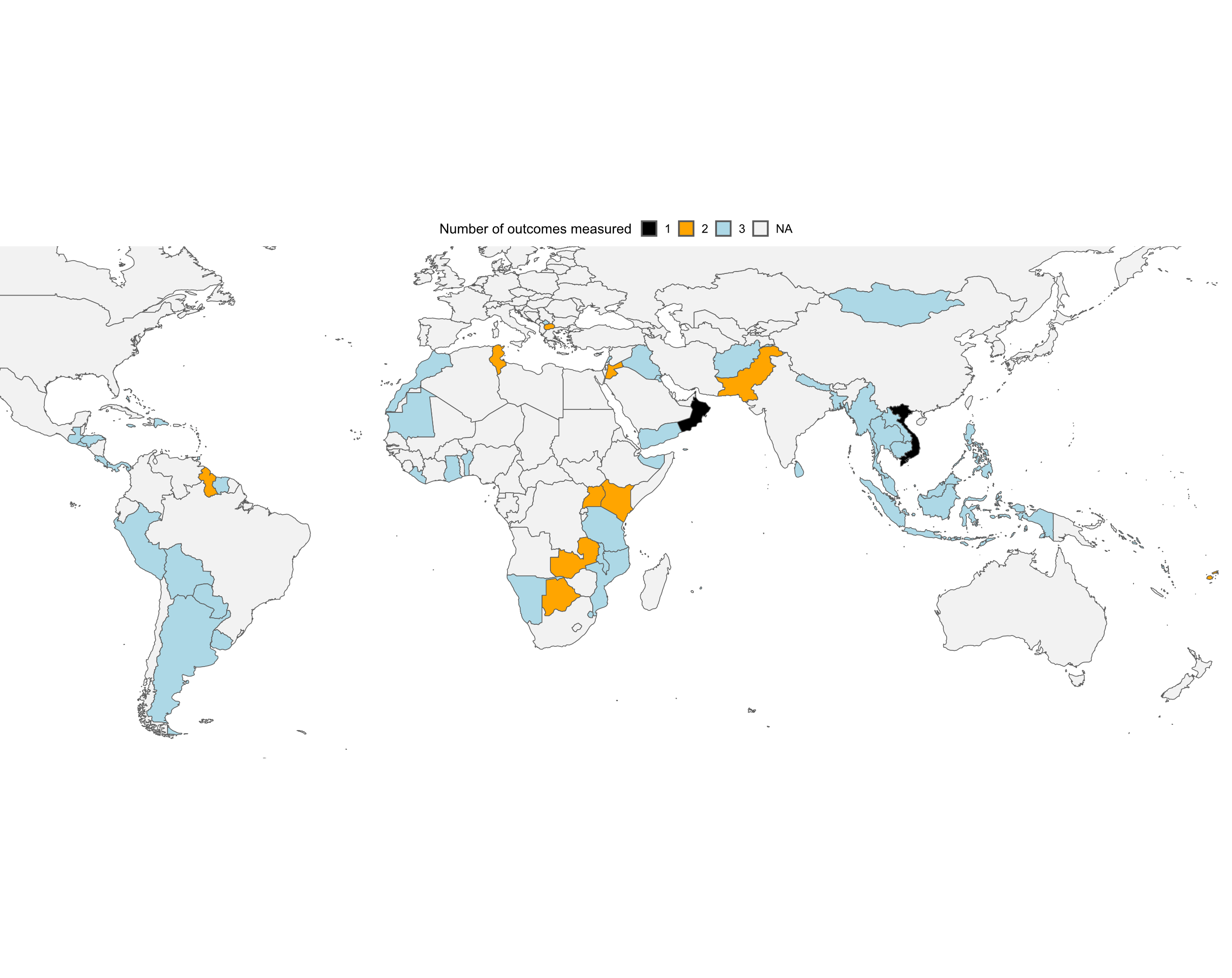

Note: Several small island countries and territories are not visible within the figure.

### **Supplementary Table 1. Country-level prevalence of suicide thoughts and behaviours and food insecurity and country-level indicator data.**

| Country | Food-insecure (%) | Reporting suicidal ideation (%) | Reporting suicide planning (%) | Reporting suicide attempt (%) | GDP per capita (PPP) | Gini index | Out-of-pocket health spending (%) | UHC Index |
| --- | --- | --- | --- | --- | --- | --- | --- | --- |
| Afghanistan | 19·8 | 18·8 | 16·3 | 13·8 | 2,069 |  | 73·1 | 28 |
| Anguilla | 5·5 | 23·4 | 22·5 | 17·3 |  |  |  |  |
| Antigua & Barbuda | 7·4 | 17·3 | 17·8 | 12·5 | 22,430 |  | 25·3 | 65 |
| Argentina | 2 | 21·5 | 17 | 15 | 23,291 | 41·3 | 27·7 | 74 |
| Bahamas | 6·9 | 18·3 | 15·5 | 13·9 | 31,856 | . | 27·9 | 64 |
| Bahrain | 11·2 | 15·2 | 14·2 | 13·1 | 44,769 | . | 28 | 72 |
| Bangladesh | 13·1 | 4·9 | 7·3 | 6·7 | 3,368 | 32·1 | 69·9 | 37 |
| Belize | 7·2 | 14·2 | 16·7 | 12·7 | 7,619 | . | 24·5 | 64 |
| Benin | 18·4 | 13·8 | 15·6 | 15·6 | 3,005 | 47·8 | 43 | 37 |
| Bolivia | 4·1 | 22·1 | 19·5 | 21·5 | 8,862 | 42·6 | 22 | 65 |
| Botswana | 14 | 23·1 | 29·4 |  | 10,345 | 64·7 | 8·8 | 46 |
| British Virgin Islands | 6·4 | 14·7 | 16·2 | 12·1 |  |  |  |  |
| Brunei Darussalam | 6·7 | 9·3 | 6·6 | 5·9 | 81,371 |  | 7·3 | 73 |
| Cambodia | 6·3 | 6·4 | 8·8 | 6·8 | 3,046 |  | 61·2 | 53 |
| Cayman Islands | 5·1 | 18·1 | 15 |  | 71,483 |  |  |  |
| Cook Islands | 10 | 15·3 | 14·6 | 13·5 |  |  |  |  |
| Costa Rica | 1·4 | 10·8 | 7·2 | 8·3 | 12,217 | 50·6 | 26·4 | 69 |
| Curacao | 3·8 | 11·3 | 9 | 10·5 | 24,221 |  |  |  |
| Dominica | 5·7 | 21·1 | 19·7 | 15 | 9,838 |  | 42·2 | 60 |
| Dominican Republic | 2·7 | 18·1 | 14·9 | 15·5 | 12,632 | 45·5 | 44·6 | 63 |
| El Salvador | 3·6 | 13·7 | 11·2 | 13 | 6,877 | 43·4 | 28·2 | 69 |
| Eswatini | 8·6 | 17 | 21 | 16·2 | 8,708 | 51·5 | 10·5 | 52 |
| Fiji | 10·7 | 17·6 | 20·9 |  | 7,915 | 40·4 | 22·4 | 56 |
| French Polynesia | 11·1 | 14 | 16·7 | 9·7 |  |  |  |  |
| Ghana | 14·6 | 19·9 | 23·2 | 26·4 | 3,782 | 42·4 | 38·3 | 34 |
| Grenada | 8·1 | 22·5 | 22·3 |  | 11,565 | . | 55·2 | 61 |
| Guatemala | 3·2 | 18 | 13·6 | 16 | 8,194 | 48·3 | 55·7 | 57 |
| Guyana | 7·8 | 23·5 | 22·9 |  | 8,895 |  | 32·8 | 63 |
| Honduras | 3·9 | 19·5 | 18·9 | 17·3 | 4,033 | 53·4 | 49·9 | 57 |
| Indonesia | 4·1 | 5·2 | 5·6 | 3·9 | 10,247 | 39·7 | 43 | 51 |
| Iraq | 8·8 | 17·6 | 16·3 | 16·1 | 15,163 | 29·5 | 61·7 | 47 |
| Jamaica | 6·8 | 24·9 | 24·9 | 17·8 | 9,600 | 45·5 | 16·8 | 69 |
| Jordan | 14·1 | 18 | 17·9 |  | 9,183 | 33·9 | 36 | 59 |
| Kenya | 14·7 | 27·9 | 29·5 |  | 1,916 | 45 | 45 | 30 |
| Kiribati | 12·8 | 34·7 | 33·6 | 31·4 | 1,739 | 37 | 0·1 | 44 |
| Kuwait | 7·9 | 17·1 | 16·9 | 17·4 | 47,231 |  | 13·7 | 67 |
| Lao | 1·2 | 2·9 | 4·3 | 5·1 | 6,168 | 36 | 45·4 | 45 |
| Lebanon | 3·3 | 13·5 | 8·5 | 9·7 | 15,954 | 31·8 | 33·1 | 69 |
| Liberia | 16·6 | 26·3 | 35·5 | 32·9 | 1,564 | 35·3 | 48 | 39 |
| Malawi | 18·5 | 12·9 | 19·8 | 10·8 | 1,380 | 39·9 | 11·2 | 31 |
| Malaysia | 4·9 | 7·9 | 6·4 | 6·8 | 22,986 | 43·9 | 33 | 68 |
| Maldives | 5·9 | 13·3 | 18 | 12·7 | 15,993 | 38·4 | 28·1 | 62 |
| Mauritania | 10·4 | 17·8 | 15·2 | 16·7 | 4,093 | 35·7 | 62·6 | 33 |
| Mauritius | 7·5 | 16·1 | 14·5 | 12·8 | 21,415 | 36·8 | 48·1 | 64 |
| Mongolia | 1·9 | 23 | 14·7 | 9·9 | 10,550 | 33·8 | 37·3 | 54 |
| Morocco | 9·2 | 16 | 13·7 | 13·2 | 7,113 | 39·5 | 54·3 | 70 |
| Mozambique | 11·3 | 18·6 | 19·2 | 19 | 1,291 | 54 | 11·3 | 43 |
| Myanmar | 2·5 | 8·9 | 6·4 | 8·5 | 4,031 | 38·1 | 76·7 | 54 |
| Namibia | 9·9 | 19·4 | 25·2 | 25·6 | 9,736 | 61 | 9·9 | 54 |
| Nauru | 14 | 28·7 | 23·4 |  | 6,662 |  | 1·9 | 47 |
| Nepal | 4·7 | 13·9 | 13·8 | 10·2 | 2,996 | 32·8 | 59·4 | 45 |
| Niue | 11·9 | 10·2 | 10·1 | 9·5 |  |  |  |  |
| North Macedonia | 1·9 | 8·6 | 5·6 |  | 9,351 |  | 37·3 | 57 |
| Oman | 4·9 | 21·9 |  |  | 35,804 |  | 5·7 | 67 |
| Pakistan | 5·8 | 7·2 | 7·6 |  | 4,353 | 29·7 | 72·3 | 32 |
| Panama | 2·3 | 18·8 | 15·2 | 14·3 | 31,781 | 49·2 | 28·7 | 76 |
| Paraguay | 2·5 | 13·8 | 12·9 | 10·7 | 12,591 | 48·5 | 43·8 | 60 |
| Peru | 3·2 | 20 | 15·3 | 17·3 | 9,731 | 45·5 | 39·5 | 72 |
| Philippines | 7·5 | 11·3 | 10·6 | 16·2 | 7,187 | 44·6 | 51·2 | 49 |
| Samoa | 12·5 | 22·9 | 22·3 | 22·1 | 6,486 | 38·7 | 11·3 | 43 |
| Seychelles | 12·5 | 21·4 | 21·9 | 19·8 | 24,740 | 46·8 | 28·9 | 66 |
| Solomon Islands | 10·9 | 27·7 | 26 | 33·5 | 2,324 | 46·1 | 2·4 | 45 |
| Sri Lanka | 3·2 | 9·4 | 6·3 | 6·6 | 12,224 | 39·3 | 50·1 | 60 |
| St Kitts & Nevis | 4·8 | 16·3 | 16·2 | 13·4 | 22,642 |  | 55·2 | 68 |
| St Lucia | 9·5 | 24·1 | 20·1 | 16·2 | 15,651 | 51·2 | 49·2 | 70 |
| St Vincent & The Grenadines | 7·2 | 27 | 24·4 | 18 | 13,776 |  | 28·9 | 72 |
| Suriname | 11·8 | 16·5 | 14 | 11·6 | 15,062 |  | 21·8 | 67 |
| Tanzania | 6·7 | 13·9 | 9·2 | 10·9 | 2,244 | 37·8 | 26 | 37 |
| Thailand | 3·9 | 12·2 | 13·7 | 12·9 | 15,822 | 36 | 12·5 | 63 |
| Timor-Leste | 11·7 | 9·6 | 9·1 | 8·6 | 2,913 | 28·7 | 7·4 | 49 |
| Tokelau | 4·4 | 24·1 | 24·5 | 27 |  |  |  |  |
| Tonga | 11·1 | 12·6 | 13·7 | 16·4 | 6,466 | 37·6 | 9·8 | 55 |
| Trinidad & Tobago | 8·1 | 22·7 | 21·8 | 13·6 | 27,278 |  | 40·8 | 72 |
| Tunisia | 8·2 | 20·8 | 14 |  | 10,095 | 37·7 | 42·8 | 57 |
| Tuvalu | 7 | 7·8 | 11·2 | 8·3 | 3,341 | 39·1 | 0·6 | 47 |
| Uganda | 1·2 | 19·6 | 21·7 |  | 1,339 | 45·2 | 45·4 | 22 |
| United Arab Emirates | 5·6 | 16·5 | 15·8 | 13·8 | 64,890 |  | 17·9 | 74 |
| Uruguay | 1·5 | 12·3 | 11·3 | 10·2 | 18,192 | 39·9 | 18·5 | 74 |
| Vanuatu | 8·1 | 15·1 | 20·9 | 22·3 | 2,918 | 37·4 | 11 | 49 |
| Vietnam | 1·1 | 16·9 |  |  | 6,689 | 35·6 | 41·3 | 59 |
| Wallis & Futuna | 14·8 | 23·7 | 28·6 | 14·4 |  |  |  |  |
| Yemen | 10·8 | 16·7 | 14·7 | 13·3 | 3,689 | 36·7 | 76·4 | 41 |
| Zambia | 28·7 | 31·9 | 41·2 |  | 2,030 | 54·3 | 25 | 30 |

### **Supplementary Table 2. The proportion of adolescents reporting food insecurity according to outcome status in the pooled sample.**

|  | Reported being food insecure in the past 30 days |
| --- | --- |
| Pooled sample | 6·4% |
| Suicidal ideation |  |
| Reported outcome | 9·4% |
| Did not report outcome | 5·8% |
| Suicide planning |  |
| Reported outcome | 11·1% |
| Did not report outcome | 6·3% |
| Suicide attempt |  |
| Reported outcome | 11·5% |
| Did not report outcome | 6·1% |

### **Supplementary Table 3. The proportion of adolescents reporting suicide thoughts and behaviours according to food insecurity status in the pooled sample.**

|  | Reported suicidal ideation in the previous 12 months | Reported suicide planning in the previous 12 months | Reported a suicide attempt in the previous 12 months |
| --- | --- | --- | --- |
| Pooled sample | 12·6% | 11·7% | 11·3% |
| Food-secure | 12·2% | 11·1% | 10·7% |
| Food-insecure | 18·8% | 18·9% | 19·4% |

### **Supplementary Table 4. Correlations between suicide thoughts and behaviours in the pooled sample.**

|  | Suicide ideation | Suicide planning |
| --- | --- | --- |
| Suicide planning | 0·57 |  |
| Suicide attempt | 0·49 | 0·48 |

### **Supplementary Table 5. Correlations between country-level indicators.**

|  | GDP per capita | Gini index | Universal Health Coverage index | Out-of-pocket health spending |
| --- | --- | --- | --- | --- |
| Gini index | 0·11 |  |  |  |
| Universal Health Coverage index | 0·61 | 0·11 |  |  |
| Out-of-pocket health spending | -0·24 | -0·3 | -0·21 |  |
| Food insecurity prevalence in school-going adolescents | -0·23 | 0·14 | -0·53 | -0·12 |

### **Supplementary Figure 3. Prevalence Ratios for suicidal ideation, suicide planning, and suicide attempts comparing food-insecure adolescents with food-secure adolescents across countries and grouped according to WHO region.**

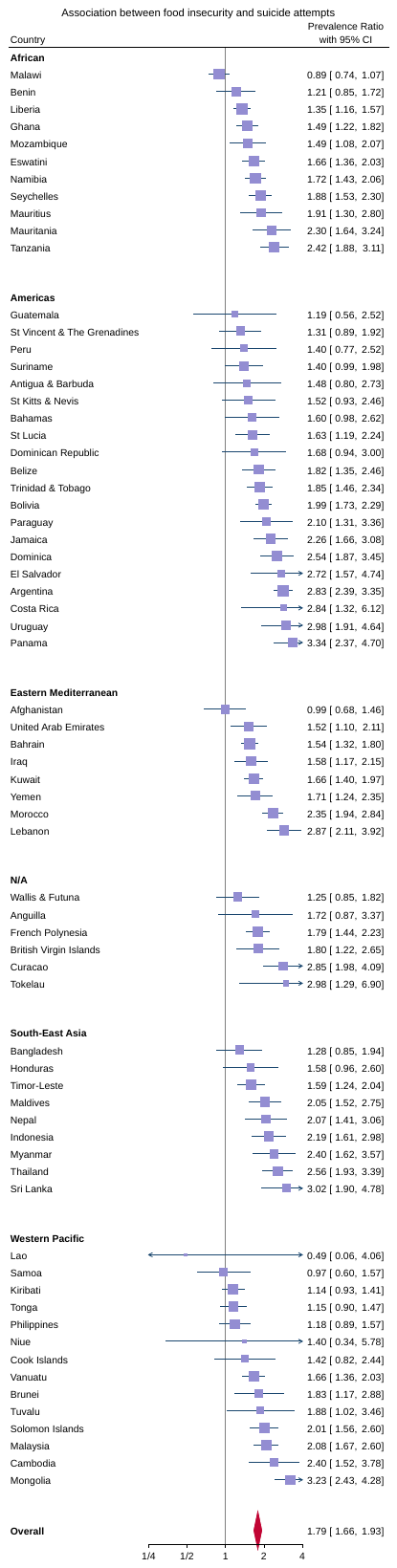

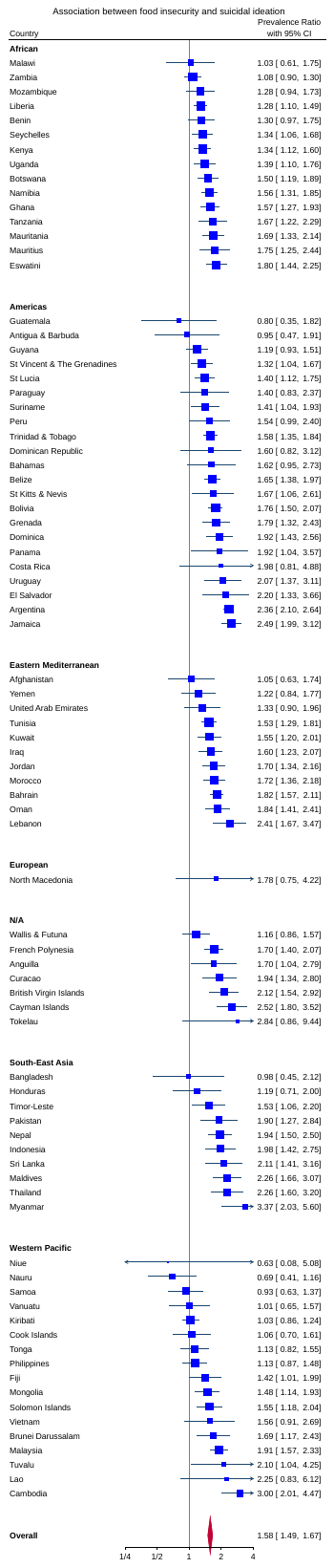

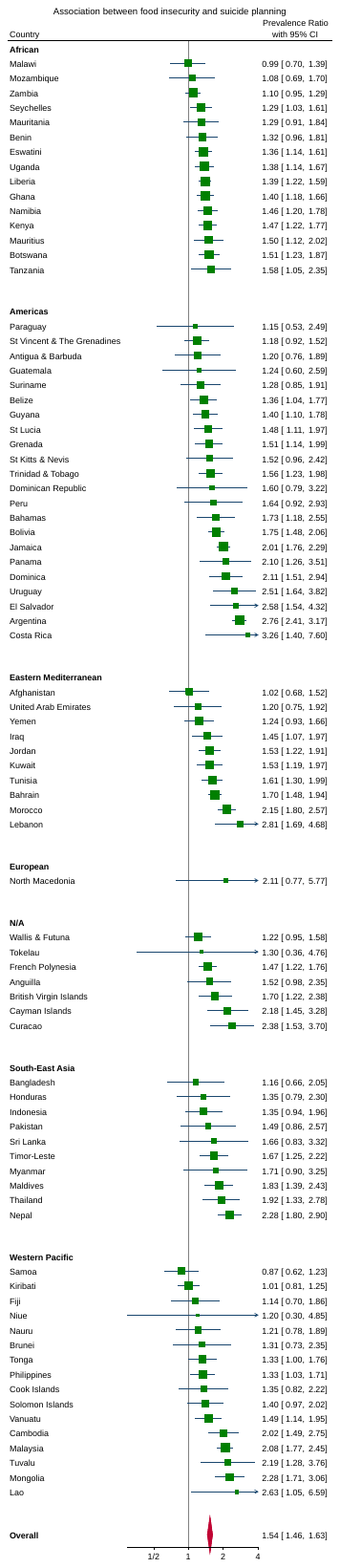

Prevalence Ratios are plotted on a log axis. The vertical line at 1 on the x-axis represents no difference in outcome prevalence between food-insecure and food-secure adolescents. Larger numbers away from 1 indicate that suicidal ideation is comparatively more prevalent in food-insecure adolescents. Smaller numbers closer to zero indicate that the outcome is comparatively more prevalent in food-secure adolescents.

### **Supplementary Table 6. Meta-regression analyses of country-level indicators associated with the magnitude of country-level prevalence ratios.**

|  | Prevalence of food insecurity | | | GDP per capita | | | Gini index | | | Universal Health Coverage index | | | Out-of-pocket health spending | | |
| --- | --- | --- | --- | --- | --- | --- | --- | --- | --- | --- | --- | --- | --- | --- | --- |
|  | Countries | Change in log-prevalence ratio per 1 percentage point increase in food insecurity prevalence | Between-country variance explained (%) | Countries | Change in log-prevalence ratio per increase in log dollars in GDP per capita | Between-country variance explained (%) | Countries | Change in log-prevalence ratio per 1-point increase in the Gini index | Between-country variance explained (%) | Countries | Change in log- prevalence ratio per 1-point increase in the UHC index | Between-country variance explained (%) | Countries | Change in log- prevalence ratio per 1 percentage point increase in out-of-pocket health spending | Between-country variance explained (%) |
| Suicidal ideation | 83 | -0·0299  (-0·0388 to -0·021) | 56·5 | 76 | 0·0961 (0·0461 to 0·148) | 28·4 | 55 | -0·00578  (-0·0146 to 0·00308) | 1 | 74 | 0·00737 (0·00361 to 0·0111) | 30·3 | 74 | 0·00205  (-0·00106 to 0·00515) | 0·1 |
| Suicide planning | 81 | -0·0307  (-0·0397 to -0·0218) | 54·2 | 74 | 0·0918 (0·0356 to 0·148) | 20·1 | 54 | -0·00583  (-0·0153 to 0·0036) | 1·5 | 72 | 0·00851 (0·00478 to 0·0122) | 35·3 | 72 | 0·00119  (-0·00199 to 0·00437) | 0·5 |
| Suicide attempt | 68 | -0·0522  (-0·0632 to -0·0411) | 77·8 | 61 | 0·1045 (0·03 to 0·179) | 15·6 | 46 | -0·00242  (-0·0165 to 0·01136) | 0 | 60 | 0·0106 (0·00516 to 0·016) | 27·6 | 60 | 0·00127  (-0·00278 to 0·00533) | 0 |

Coefficients and 95% confidence intervals are presented. A positive coefficient indicates that an increase in the country-level indicator is associated with an increase in the magnitude of the association between food insecurity and the outcome of interest. A negative coefficient indicates that an increase in the country-level indicator is associated with a decrease in the magnitude of the association between food insecurity and the outcome of interest.

### **Supplementary Table 7. Adjusted meta-regression analyses of country-level indicators associated with the magnitude of country-level prevalence ratios.**

|  | Prevalence of food insecurity^a^ | | | Gini index^b^ | | | Universal Health Coverage index^b^ | | | Out-of-pocket health spending^b^ | | |
| --- | --- | --- | --- | --- | --- | --- | --- | --- | --- | --- | --- | --- |
|  | Countries | Change in log-prevalence ratio per 1 percentage point increase in food insecurity prevalence | Between-country variance explained (%) | Countries | Change in log-prevalence ratio per 1-point increase in the Gini index | Between-country variance explained (%) | Countries | Change in log- prevalence ratio per 1-point increase in the UHC index | Between-country variance explained (%) | Countries | Change in log- prevalence ratio per 1 percentage point increase in out-of-pocket health spending | Between-country variance explained (%) |
| Suicidal ideation | 55 | -0·0209  (-0·0329 to -0·009) | 64·6 | 55 | -0·00605  (-0·0134 to 0·00132) | 44·6 | 74 | 0·00734 (0·000426 to 0·0142) | 28·1 | 74 | 0·00268  (-0·000197 to 0·00556) | 26·6 |
| Suicide planning | 54 | -0·025  (-0·038 to -0·0119) | 57·5 | 54 | -0·00717  (-0·0153 to 0·000937) | 37·8 | 72 | 0·0121  (0·00523 to 0·0191) | 35·1 | 72 | 0·00176  (-0·00125 to 0·00476) | 19·4 |
| Suicide attempt | 46 | -0·0458  (-0·0631 to -0·0286) | 75·5 | 46 | -0·00667  (-0·0183 to 0·00499) | 40·3 | 60 | 0·00175  (-0·00209 to 0·00558) | 26·2 | 60 | 0·0138  (0·00389 to 0·0237) | 14·7 |

^a^adjusted for GDP per capita (PPP) and Gini index. ^b^adjusted for GDP per Capita (PPP). Coefficients and 95% confidence intervals are presented. A positive coefficient indicates that an increase in the country-level indicator is associated with an increase in the magnitude of the association between food insecurity and the outcome of interest. A negative coefficient indicates that an increase in the country-level indicator is associated with a decrease in the magnitude of the association between food insecurity and the outcome of interest.

### **Supplementary Table 8. Poisson regression coefficients with 95% confidence intervals of the mixed-effects prevalence ratio analyses.**

|  | Suicidal ideation | Suicide planning | Suicide attempt |
| --- | --- | --- | --- |
| Model A: Food insecurity only  Food-insecure | 1·58 (1·49 to 1·66) | 1·56 (1·48 to 1·64) | 1·79 (1·68 to 1·91) |
| Model B: Individual-level confounders  Food-insecure | 1·59 (1·5 to 1·68) | 1·57 (1·49 to 1·65) | 1·8 (1·69 to 1·93) |
| Model C: Prevalence of food insecurity  Food-insecure  Percent food-insecure | 1·58 (1·5 to 1·67)  1·05 (0·95 to 1·17) | 1·54 (1·46 to 1·62)  1·22 (1·09 to 1·35) | 1·79 (1·68 to 1·92)  1·09 (0·96 to 1·24) |
| Model D: Prevalence of food insecurity interaction  Food-insecure  Percent food-insecure  Percent food-insecure X Food-insecure | 1·63 (1·56 to 1·71)  1·14 (1·05 to 1·24)  0·87 (0·84 to 0·91) | 1·61 (1·54 to 1·68)  1·27 (1·16 to 1·38)  0·88 (0·84 to 0·91) | 1·82 (1·73 to 1·92)  1·17 (1·05 to 1·3)  0·82 (0·76 to 0·88) |
| Model E: Prevalence of food insecurity interaction adjusted^a^  Food-insecure  Percent food-insecure  Percent food-insecure x Food-insecure | 1·63 (1·55 to 1·72)  1·16 (1·03 to 1·31)  0·88 (0·84 to 0·92) | 1·62 (1·54 to 1·71)  1·27 (1·14 to 1·341)  0·88 (0·84 to 0·92) | 1·84 (1·72 to 1·96)  1·25 (1·108 to 1·45)  0·82 (0·76 to 0·88) |
| Model F: GDP (log)  Food-insecure  GDP per capita | 1·6 (1·51 to 1·69)  1·07 (0·98 to 1·17) | 1·57 (1·49 to 1·66)  0·97 (0·88 to 1·06) | 1·82 (1·69 to 1·96)  0·95 (0·86 to 1·06) |
| Model G: GDP (log) interaction  Food-insecure  GDP per capita  GDP per capita X Food-insecure | 1·61 (1·53 to 1·69)  1 (0·91to 1·1)  1·08 (1·03 to 1·14) | 1·59 (1·51 to 1·67)  0·91 (0·83 to 1·01)  1·08 (1·02 to 1·13) | 1·83 (1·71 to 1·96)  0·91 (0·82 to 1·01)  1·09 (1·02 to 1·17) |
| Model H: Gini  Food-insecure  Gini | 1·59 (1·49 to 1·7)  1·16 (1·04 to 1·29) | 1·58 (1·48 to 1·69)  1·19 (1·07 to 1·32) | 1·85 (1·7 to 2·02)  1·18 (1·03 to 1·34) |
| Model I: Gini interaction  Food-insecure  Gini  Gini X Food-insecure | 1·59 (1·49 to 1·7)  1·19 (1·07 to 1·34)  0·95 (0·9 to 1·02) | 1·58 (1·48 to 1·69)  1·24 (1·11 to 1·38)  0·96 (0·9 to 1·02) | 1·85 (1·7 to 2·01)  1·19 (1·04 to 1·37)  0·98 (0·89 to 1·07) |
| Model J: Gini interaction adjusted^b^  Food-insecure  Gini  Gini X Food-insecure | 1·59 (1·49 to 1·7)  1·19 (1·07 to 1·33)  0·95 (0·9 to 1·02) | 1·58 (1·48 to 1·68)  1·24 (1·11 to 1·38)  0·96 (0·9 to 1·02) | 1·84 (1·69 to 2·01)  1·21 (1·06 to 1·38)  0·98 (0·89 to 1·07) |
| Model K: UHC index  Food-insecure  UHC index | 1·6 (1·51 to 1·69)  1·12 (1·02 to 1·23) | 1·57 (1·49 to 1·66)  1·02 (0·92 to 1·12) | 1·82 (1·69 to 1·95)  1·02 (0·91 to 1·14) |
| Model L: UHC index interaction  Food-insecure  UHC index  UHC index X Food-insecure | 1·6 (1·52 to 1·68)  1·03 (0·93 to 1·14)  1·09 (1·04 to 1·14) | 1·57 (1·5 to 1·65)  0·94 (0·84 to 1·04)  1·1 (1·05 to 1·15) | 1·8 (1·68 to 1·92)  0·95 (0·85 to 1·07)  1·13 (1·05 to 1·2) |
| Model M: UHC index adjusted^b^  Food-insecure  UHC index  UHC index X Food-insecure | 1·6 (1·52 to 1·68)  1·1 (0·96 to 1·27)  1·09 (1·04 to 1·15) | 1·58 (1·5 to 1·66)  1·06 (0·91 to 1·23)  1·1 (1·05 to 1·15) | 1·8 (1·69 to 1·92)  1·11 (0·94 to 1·23)  1·12 (1·05 to 1·2) |
| Model N: OOP health spending  Food-insecure  OOP health spending | 1·58 (1·5 to 1·68)  0·91 (0·83 to 0·99) | 1·56 (1·48 to 1·65)  0·88 (0·8 to 0·96) | 1·81 (1·69 to 1·95)  0·89 (0·8 to 0·98) |
| Model O: OOP health spending interaction  Food-insecure  OOP health spending  OOP health spending X Food-insecure | 1·59 (1·5 to 1·69)  0·9 (0·81 to 0·99)  1·02 (0·96 to 1·08) | 1·56 (1·48 to 1·65)  0·87 (0·78 to 0·96)  1·02 (0·97 to 1·08) | 1·81 (1·69 to 1·95)  0·89 (0·8 to 0·99)  0·99 (0·93 to 1·07) |
| Model P: OOP health spending adjusted^b^  Food-insecure  OOP health spending  OOP health spending X Food-insecure | 1·59 (1·5 to 1·68)  0·9 (0·82 to 1)  1·02 (0·96 to 1·08) | 1·56 (1·48 to 1·65)  0·86 (0·78 to 0·95)  1·02 (0·97 to 1·08) | 1·81 (1·68 to 1·95)  0·88 (0·8 to 0·98)  0·99 (0·93 to 1·07) |

^a^adjusted for log GDP per capita (PPP) and Gini index. ^b^adjusted for GDP per Capita

### **Supplementary Table** **9.** **Poisson regression coefficients with 95% confidence intervals of the mixed-effects Relative Index of Inequality analyses.**

|  | Suicidal ideation | Suicide planning | Suicide attempt |
| --- | --- | --- | --- |
| Model A: Food insecurity only  RII | 0·49 (0·46 to 0·53) | 0·53 (0·5 0 to 0·57) | 0·44 (0·41 to 0·47) |
| Model B: Individual-level confounders  RII | 0·49 (0·46 to 0·53) | 0·53 (0·49 to 0·56) | 0·43 (0·4 to 0·47) |
| Model C: GDP (log)  RII  GDP per capita | 0·49 (0·46 to 0·52)  1·02 (0·93 to 1·13) | 0·53 (0·49 to 0·56)  0·92 (0·83 to 1·02) | 0·43 (0·4 to 0·47)  0·92 (0·82 to 1·02) |
| Model D: GDP (log) interaction  RII  GDP per capita  GDP per capita X RII | 0.49 (0·46 to 0·352)  1·04 (0·94 to 1·14)  0·92 (0·86 to 0·99) | 0·53 (0·49 to 0·256)  0·93 (0·84 to 1·03)  0·95 (0·89 to 1·01) | 0·43 (0·4 to 0·46)  0·93 (0·83 to 1·04)  0·94 (0·88 to 1.02) |
| Model E: Gini  RII  Gini | 0·49 (0·45 to 0·53)  1·19 (1·07 to 1·33) | 0·52 (0·48 to 0·56)  1·24 (1·1 to 1·39) | 0·42 (0·38 to 0·45)  1·19 (1·04 to 1·37) |
| Model F: Gini interaction  RII  Gini  Gini X RII | 0·49 (0·45 to 0·53)  1·2 (1·06 to 1·34)  0·99 (0·91 to 1·08) | 0·52 (0·48 to 0·56)  1·24 (1·1 to 1·4)  0·98 (0·9 to 1·05) | 0·42 (0·38 to 0·45)  1·19 (1·04 to 1·37)  0·99 (0·9 to 1·1) |
| Model G: Gini interaction adjusted^a^  RII  Gini  Gini X RII | 0·49 (0·45 to 0·53)  1·2 (1·07 to 1·35)  0·99 (0·91 to 1·08) | 0·52 (0·48 to 0·56)  1·26 (1·12 to 1·41)  0·97 (0·9 to 1·05) | 0·42 (0·38 to 0·46)  1·22 (1·06 to 1·4)  0·99 (0·9 to 1·09) |
| Model H: UHC index  RII  UHC index | 0·49 (0·46 to 0·53)  1·07 (0·96 to 1·19) | 0·53 (0·5 to 0·57)  0·95 (0·85 to 1·06) | 0·43 (0·4 to 0·47)  0·97 (0·86 to 1·1) |
| Model I: UHC index interaction  RII  UHC index  UHC index X RII | 0·5 (0·47 to 0·53)  1·08 (0·98 to 1·19)  0·9 (0·84 to 0·96) | 0·53 (0·5 to 0·57)  0·97 (0·87 to 1·08)  0·92 (0·87 to 0·98) | 0·44 (0·41 to 0·47)  0·99 (0·89 to 1·11)  0·9 (0·83 to 0·97) |
| Model J: UHC index adjusted^a^  RII  UHC index  UHC index X RII | 0·5 (0·47 to 0·53)  1·17 (1·01 to 1·37)  0·9 (0·84 to 0·96) | 0·53 (0·5 to 0·57)  1·08 (0·92 to 1·27)  0·92 (0·87 to 0·98) | 0·44 (0·41 to 0·47)  1·17 (0·98 to 1·4)  0·9 (0·83 to 0·97) |
| Model K: OOP health spending  RII  OOP health spending | 0·5 (0·46 to 0·53)  0·91 (0·82 to 1) | 0·53 (0·5 to 0·57)  0·87 (0·79 to 0·97) | 0·43 (0·4 to 0·47)  0·89 (0·8 to 0·99) |
| Model L: OOP health spending interaction  RII  OOP health spending  OOP health spending X RII | 0·5 (0·46 to 0·53)  0·91 (0·83 to 1·01)  0·96 (0·89 to 1·03) | 0·53 (0·5 to 0·57)  0·88 (0·8 to 0·98)  0·96 (0·89 to 1·02) | 0·43 (0·4 to 0·47)  0·9 (0·81 to 1)  0·96 (0·89 to 1·04) |
| Model M: OOP health spending adjusted^a^  RII  OOP health spending  OOP health spending X RII | 0·5 (0·46 to 0·53)  0·91 (0·83 to 1·01)  0·96 (0·89 to 1·03) | 0·53 (0·5 to 0·57)  0·87 (0·79 to 0·96)  0·96 (0·89 to 1·02) | 0·43 (0·4 to 0·47)  0·89 (0·8 to 0·99)  0·96 (0·89 to 1·04) |

^a^adjusted for log GDP per capita (PPP).

### **Supplementary Table 10. Meta-regression analyses of country-level indicators associated with the magnitude of country-level Relative Index of Inequality for the 68 countries where all three outcomes were measured.**

|  | GDP per capita | | | Gini index | | | Universal Health Coverage index | | | Out-of-pocket health spending | | |
| --- | --- | --- | --- | --- | --- | --- | --- | --- | --- | --- | --- | --- |
|  | N | Change in RII per increase in log dollars in GDP per capita | Between-country variance explained (%) | N | Change in RII per 1-point increase in the Gini index | Between-country variance explained (%) | N | Change in RII per 1-point increase in the UHC index | Between-country variance explained (%) | N | Change in RII per 1 percentage point increase in out-of-pocket health spending | Between-country variance explained (%) |
| Suicidal ideation |  |  |  |  |  |  |  |  |  |  |  |  |
| Unadjusted | 61 | -0·091  (-0·182 to -0·00005) | 6·6 | 46 | -0·00907  (-0·025 to 0·00684) | 0 | 60 | -0·0104  (-0·017 to -0·00391) | 17·2 | 60 | -0·00213  (-0·00682 to 0·00256) | 0 |
| Adjusted^a^ |  |  |  | 46 | -0·006  (-0·0212 to 0·00923) | 14·2 | 60 | -0·0157  (-0·027 to -0·00443) | 16 | 60 | -0·00262  (-0·00723 to 0·00199) | 6·8 |
| Suicide planning |  |  |  |  |  |  |  |  |  |  |  |  |
| Unadjusted | 61 | -0·0668  (-0·148 to 0·0148) | 2·4 | 46 | -0·00705  (-0·0207 to 0·0066) | 0 | 60 | -0·00216  (-0·00626 to 0·00194) | 20·9 | 60 | -0·00962  (-0·0154 to -0·00384) | 1 |
| Adjusted^a^ |  |  |  | 46 | -0·00432  (-0·0174 to 0·0088) | 13·3 | 60 | -0·0172  (-0·0268 to -0·00762) | 26·9 | 60 | -0·00258  (-0·00667 to 0·00151) | 4·2 |

^a^adjusted for GDP per capita (PPP). Coefficients and 95% confidence intervals are presented. A positive coefficient indicates that an increase in the country-level indicator is associated with a reduced magnitude of inequalities in suicidal thoughts and behaviours according to food insecurity. A negative coefficient indicates that an increase in the country-level indicator is associated with an increased magnitude of inequalities in suicidal thoughts and behaviours according to food insecurity.

### **Supplementary Table 11. Unadjusted meta-regression analyses of country-level indicators associated with the magnitude of country-level prevalence ratios for the 68 countries where all three outcomes were measured.**

|  | Prevalence of food insecurity | | | GDP per capita | | | Gini index | | | Universal Health Coverage index | | | Out-of-pocket health spending | | |
| --- | --- | --- | --- | --- | --- | --- | --- | --- | --- | --- | --- | --- | --- | --- | --- |
|  | N | Change in log-prevalence ratio per 1 percentage point increase in food insecurity prevalence | Between-country variance explained (%) | N | Change in log-prevalence ratio per increase in log dollars in GDP per capita | Between-country variance explained (%) | N | Change in log-prevalence ratio per 1-point increase in the Gini index | Between-country variance explained (%) | N | Change in log- prevalence ratio per 1-point increase in the UHC index | Between-country variance explained (%) | N | Change in log- prevalence ratio per 1 percentage point increase in out-of-pocket health spending | Between-country variance explained (%) |
| Suicidal ideation | 68 | -0·0366  (-0·0486 to -0·0246) | 57·8 | 61 | 0·0777 (0·015 to 0·14) | 16·7 | 46 | -0·00366  (-0·0153 to 0·008) | 0 | 60 | 0·00773 (0·00302 to 0·0124) | 23·4 | 60 | 0·00151  (-0·00197 to 0·005) | 0 |
| Suicide planning | 68 | -0·0411  (-0·053 to -0·0292) | 59·8 | 61 | 0·0832 (0·0153 to 0·151) | 13·2 | 46 | -0·00541  (-0·0178 to 0·00695) | 0 | 60 | 0·00982  (-0·00509 to 0·0145) | 33·5 | 60 | 0·000937  (-0·00268 to 0·00455) | 0 |

A positive coefficient indicates that an increase in the country-level indicator is associated with an increase in the magnitude of the association between food insecurity and the outcome of interest. A negative coefficient indicates that an increase in the country-level indicator is associated with a decrease in the magnitude of the association between food insecurity and the outcome of interest.

### **Supplementary Table 12. Adjusted meta-regression analyses of country-level indicators associated with the magnitude of country-level prevalence ratios for the 68 countries where all three outcomes were measured.**

|  | Prevalence of food insecurity^a^ | | | Gini index^b^ | | | Universal Health Coverage index^b^ | | | Out-of-pocket health spending^b^ | | |
| --- | --- | --- | --- | --- | --- | --- | --- | --- | --- | --- | --- | --- |
|  | N | Change in log-prevalence ratio per 1 percentage point increase in food insecurity prevalence | Between-country variance explained (%) | N | Change in log-prevalence ratio per 1-point increase in the Gini index | Between-country variance explained (%) | N | Change in log- prevalence ratio per 1-point increase in the UHC index | Between-country variance explained (%) | N | Change in log- prevalence ratio per 1 percentage point increase in out-of-pocket health spending | Between-country variance explained (%) |
| Suicidal ideation | 46 | -0·0305  (-0·048 to -0·0129) | 58·3 | 46 | -0·00569  (-0·0158 to 0·00442) | 34·4 | 60 | 0·00897  (0·00068 to 0·0172) | 20·9 | 60 | 0·00187  (-0·00145 to 0·00519) | 16 |
| Suicide planning | 46 | -0·0386  (-0·0565 to -0·0208) | 63·6 | 46 | -0·00945  (-0·0203 to 0·00145) | 34·8 | 60 | 0·015  (0·00673 to 0·0233) | 34·6 | 60 | 0·00137  (-0·0021 to 0·00484) | 12·4 |

^a^adjusted for GDP per capita (PPP) and Gini index. ^b^adjusted for GDP per Capita (PPP). A positive coefficient indicates that an increase in the country-level indicator is associated with an increase in the magnitude of the association between food insecurity and the outcome of interest. A negative coefficient indicates that an increase in the country-level indicator is associated with a decrease in the magnitude of the association between food insecurity and the outcome of interest.

### **Supplementary Table 13. Poisson regression coefficients with 95% confidence intervals of the mixed-effects prevalence ratio and Relative Index of Inequality analyses for the 68 countries where all three outcomes were measured.**

|  | Suicidal ideation | | Suicide planning | |
| --- | --- | --- | --- | --- |
|  | RII | Binary | RII | Binary |
| Model A: Food insecurity only  RII/Food-insecure | 0·49 (0·46 to 0·53) | 1·6 (1·51 to 1·69) | 0·54 (0·5 to 0·58) | 1·57 (1·48 to 1·67) |
| Model B: Individual-level confounders  RII/Food-insecure | 0·49 (0·45 to 0·52) | 1·61 (1·52 to 1·71) | 0·53 (0·5 to 0·57) | 1·58 (1·49 to 1·68) |
| Model C: Prevalence of food insecurity  RII/Food-insecure % food-insecure | - | 1·61 (1·52 to 1·71)  1·01 (0·9 to 1·14) | - | 1·57 (1·48 to 1·66)  1·15 (1·01 to 1·31) |
| Model D: Prevalence of food insecurity interaction  RII/Food-insecure  % food-insecure  % food-insecure X RII/food-insecure | - | 1·64 (1·56 to 1·72)  1·1 (0·99 to 1·23)  0·86 (0·81 to 0·91) | - | 1·61 (1·54 to 1·69)  1·23 (1·1 to 1·36)  0·85 (0·8 to 0·89) |
| Model E: Prevalence of food insecurity interaction adjusted^a^  RII/Food-insecure  % food-insecure  % food-insecure x RII/food-insecure | - | 1·63 (1·53 to 1·73)  1·17 (1·01 to 1·37)  0·86 (0·81 to 0·92) | - | 1·62 (1·53 to 1·72)  1·3 (1·14 to 1·5)  0·84 (0·79 to 0·89) |
| Model F: GDP (log)  RII/Food-insecure  GDP per capita | 0·48 (0·45 to 0·52)  1·01 (0·9 to 1·13) | 1·63 (1·53 to 1·73)  1·06 (0·95 to 1·18) | 0·53 (0·49 to 0·57)  0·94 (0·84 to 1.05) | 1·59 (1·5 to 1·7)  0·98 (0·88 to 1·09) |
| Model G: GDP (log) interaction  RII/Food-insecure  GDP per capita  GDP per capita X RII/food-insecure | 0·48 (0·45 to 0·52)  1·02 (0·91 to 1·14)  0·97 (0·89 to 1·05) | 1·63 (1·54 to 1·373)  1·02 (0·91 to 1·14)  1·06 (1·01 to 1·13) | 0·85 (0·61 to 1·2)  0·96 (0·9 to 1·01)  0·99 (0·91 to 1·07) | 1·6 (1·51 to 1·7)  0·94 (0·84 to 1·04)  1·07 (1·01 to 1·13) |
| Model H: Gini  RII/Food-insecure  Gini | 0·48 (0·44 to 0·52)  1·18 (1·03 to 1·36) | 1·63 (1·51 to 1·75)  1·17 (1·02 to 1·34) | 0·52 (0·48 to 0·57)  1·19 (1·04 to 1·37) | 1·62 (1·5 to 1·74)  1·15 (1·01 to 1·31) |
| Model I: Gini interaction  RII/Food-insecure  Gini  Gini X Food-insecure | 0·48 (0·44 to 0·52)  1·21 (1·04 to 1·4)  0·95 (0·86 to 1·05) | 1·63 (1·51 to 1·75)  1·18 (1·02 to 1·36)  0·97 (0·9 to 1·05) | 0·52 (0·48 to 0·57)  1·21 (1·05 to 1·4)  0·94 (0·86 to 1·04) | 1·62 (1·5 to 1·74)  1·19 (1·04 to 1·36)  0·96 (0·88 to 1·03) |
| Model J: Gini interaction adjusted^b^  RII/Food-insecure  Gini  Gini X RII/food-insecure | 0·48 (0·44 to 0·52)  1·22 (1·05 to 1·41)  0·95 (0·86 to 1·05) | 1·63 (1·51 to 1·75)  1·17 (1·02 to 1·35)  0·97 (0·9 to 1·05) | 0·52 (0·48 to 0·57)  1·24 (1·07 to 1·43)  0·94 (0·86 to 1·04) | 1·61 (1·5 to 1·73)  1·2 (1·05 to 1·37)  0·96 (0·88 to 1·03) |
| Model K: UHC index  RII/Food-insecure  UHC index | 0·49 (0·45 to .52)  1·1 (0·98 to 1·24) | 1·63 (1·53 to 1·73)  1·16 (1·04 to 1·29) | 0·54 (0·5 to 0·58)  0·98 (0·87 to 1·11) | 1·6 (1·5 to 1·7)  1·06 ( 0·95 to 1·19) |
| Model L: UHC index interaction  RII/Food-insecure  UHC index  UHC index X RII/food-insecure | 0·5 (0·46 to 0·53)  1·13 (1·01 to 1·28)  0·89 (0·82 to 0·96) | 1·61 (1·52 to 1·71)  1·08 (0·96 to 1·21)  1·09 (1·03 to 1·15) | 0·55 (0·51 to 0·58)  1·02 (0·91 to 1·15)  0·9 (0·84 to 0·96) | 1·58 (1·49 to 1·67)  0·98 (0·87 to 1·1)  1·11 (1·05 to 1·17) |
| Model M: UHC index adjusted^b^  RII/Food-insecure  UHC index  UHC index X RII/food-insecure | 0·5 (0·46 to 0·53)  1·27 (1·07 to 1·51)  0·89 (0·82 to 0·96) | 1·62 (1·53 to 1·71)  1·2 (1·01 to 1·44)  1·09 (1·03 to 1·15) | 0·55 (0·51 to 0·58)  1·16 (0·98 to 1·38)  0·9 (0·84 to 0·97) | 1·58 (1·5 to 1·68)  1·15 (0·99 to 1·34)  1·11 (1·05 to 1·17) |
| Model N: OOP health spending  RII/Food-insecure  OOP health spending | 0·49 (0·45 to 0·53)  0·92 (0·83 to 1·02) | 1·62 (1·52 to 1·72)  0·92 (0·83 to 1·02) | 0·54 (0·5 to 0·58)  0·88 (0·79 to 0·98) | 1·59 (1·49 to 1·69)  0·89 (0·81 to 0·98) |
| Model O: OOP health spending interaction  RII/Food-insecure  OOP health spending  OOP health spending X RII/food-insecure | 0·49 (0·45 to 0·53)  0·92 (0·82 to 1·03)  0·98 (0·91 to 1·07) | 1·62 (1·52 to 1·73)  0·92 (0·82 to 1·02)  1 (0·94 to 1·07) | 0·54 (0·5 to 0·58)  0·89 (0·8 to 0·99)  0·97 (0·9 to 1·05) | 1·59 (1·49 to 1·69)  0·88 (0·79 to 0·98)  1·02 (0·96 to 1·08) |
| Model P: OOP health spending adjusted^b^  RII/Food-insecure  OOP health spending  OOP health spending X RII/food-insecure | 0·49 (0·45 to 0·53)  0·92 (0·82 to 1·03)  0·98 (0·91 to 1·06) | 1·62 (1·52 to 1·73)  0·92 (0·83 to 1·03)  1 (0·94 to 1·07) | 0·54 (0·5 to 0·58)  0·88 (0·79 to 0·99)  0·97 (0·9 to 1·05) | 1·59 (1·49 to 1·69)  0·88 (0·79 to 0·97)  1·02 (0·96 to 1·08) |

^a^adjusted for GDP per capita (PPP) and Gini index. ^b^adjusted for GDP per Capita (PPP).

### **Supplementary Figure 4. Relative Index of Inequality for suicidal ideation, suicide planning, and suicide attempts comparing the most food-secure adolescents with the least food-secure adolescents across countries with multiple surveys. Overlapping 95% confidence intervals suggest no difference in the magnitude of inequality across time within a country.**

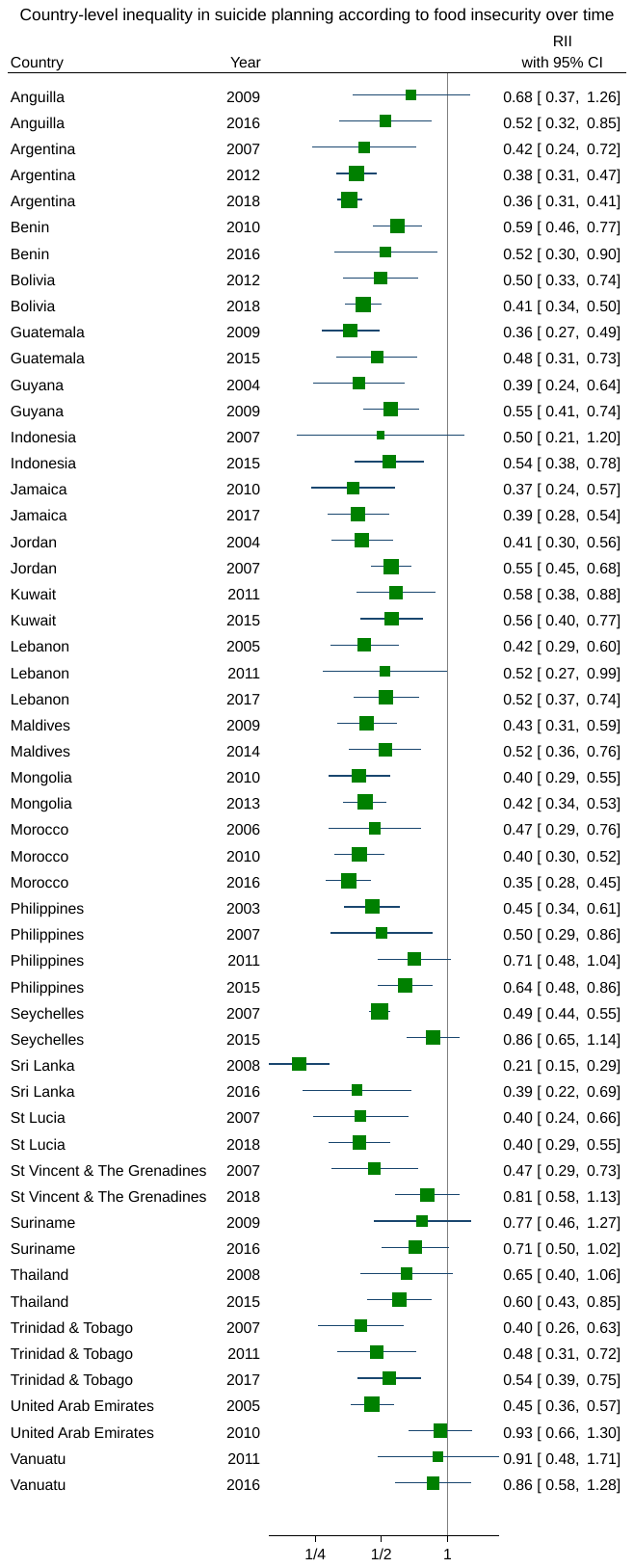

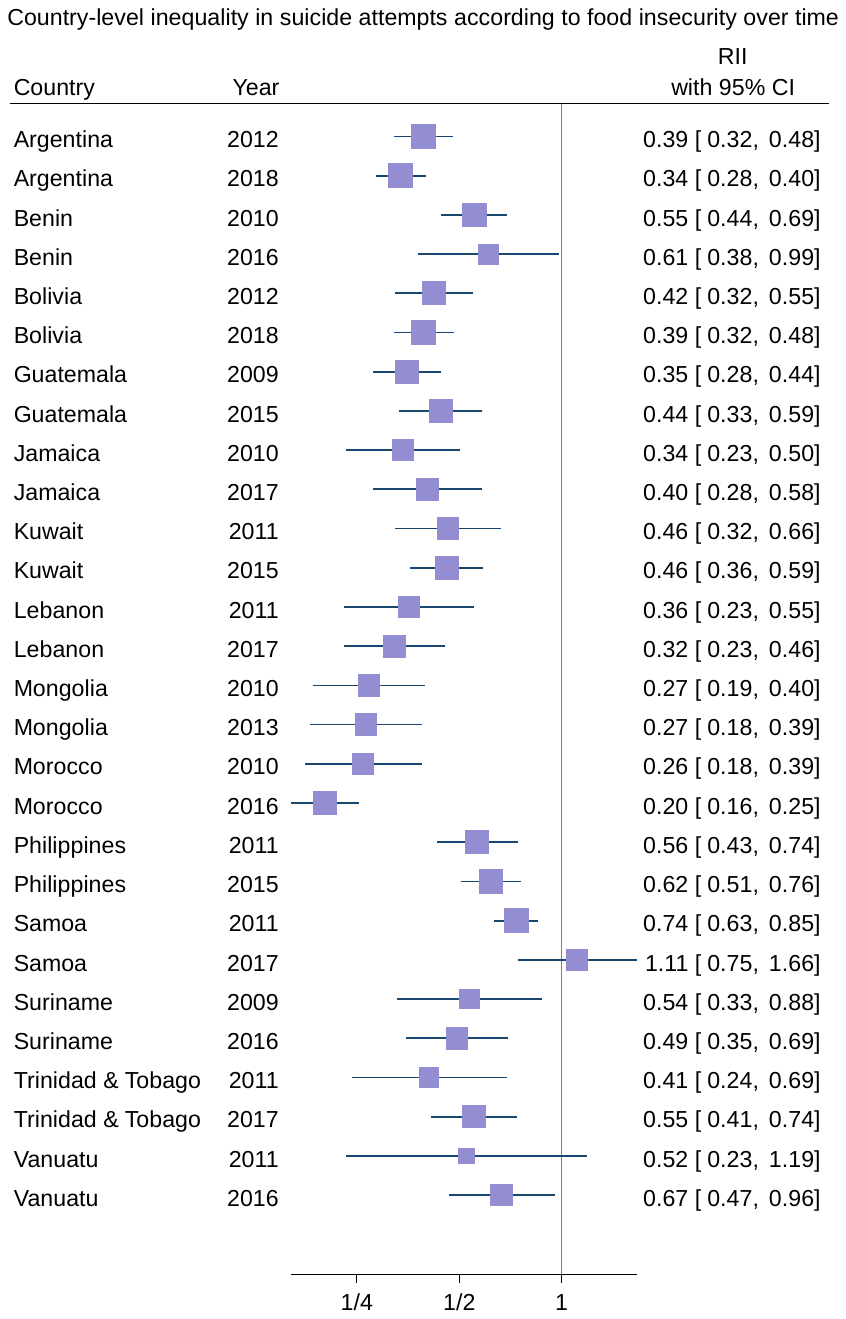

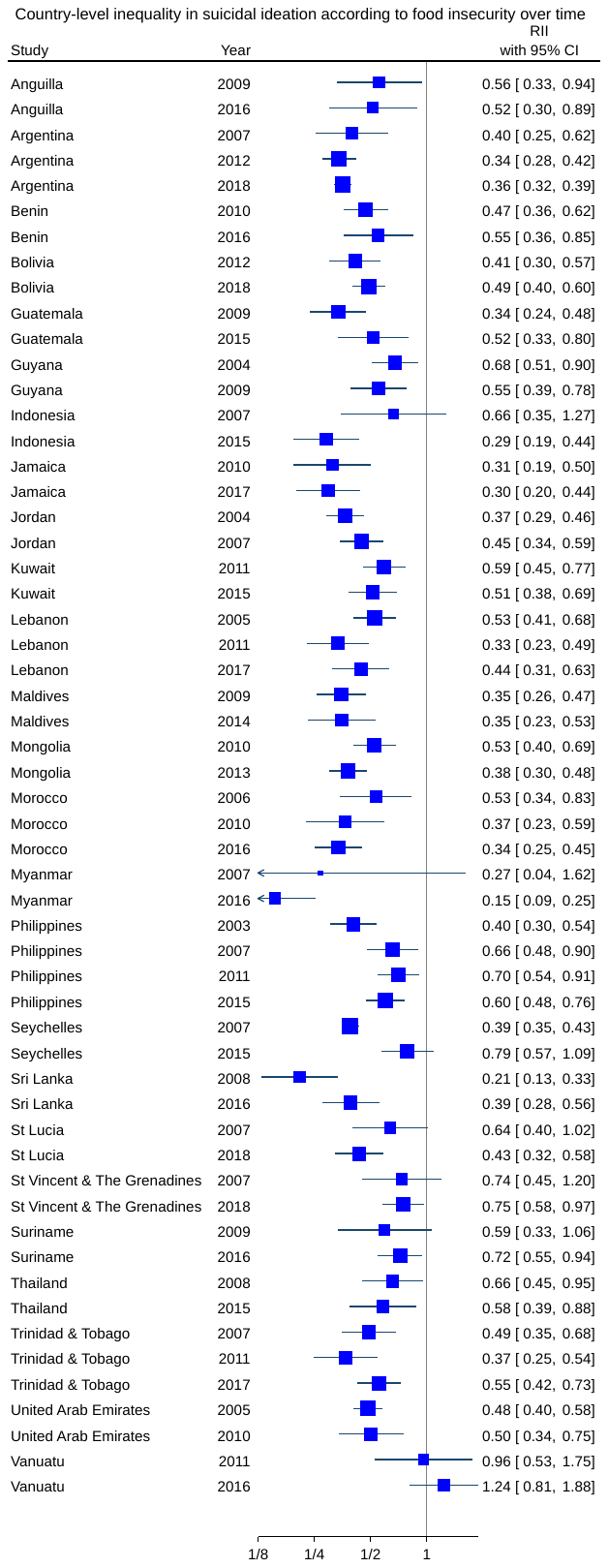

### **Supplementary Figure 5. Prevalence Ratios for suicidal ideation, suicide planning, and suicide attempts comparing the most food-insecure adolescents with food-secure adolescents across countries with multiple surveys. Overlapping 95% confidence intervals suggest no difference in the magnitude of the association across time within a country.**

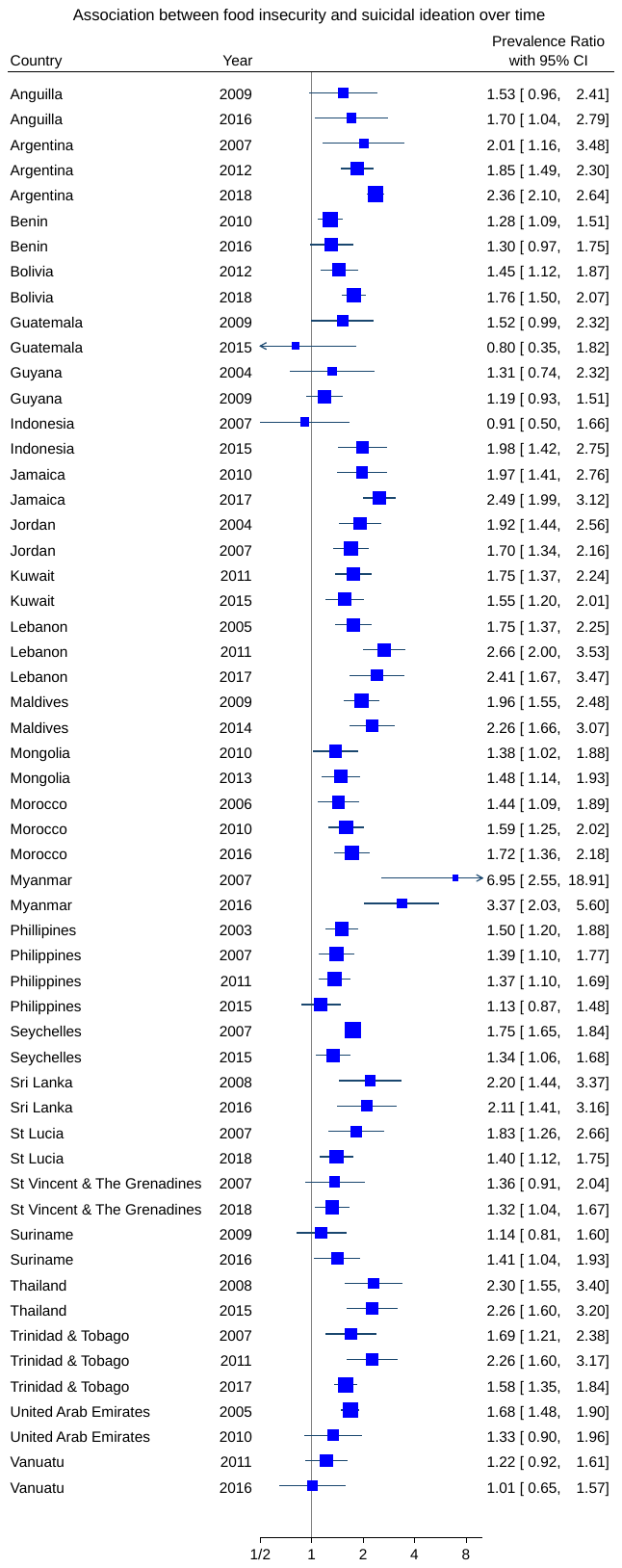

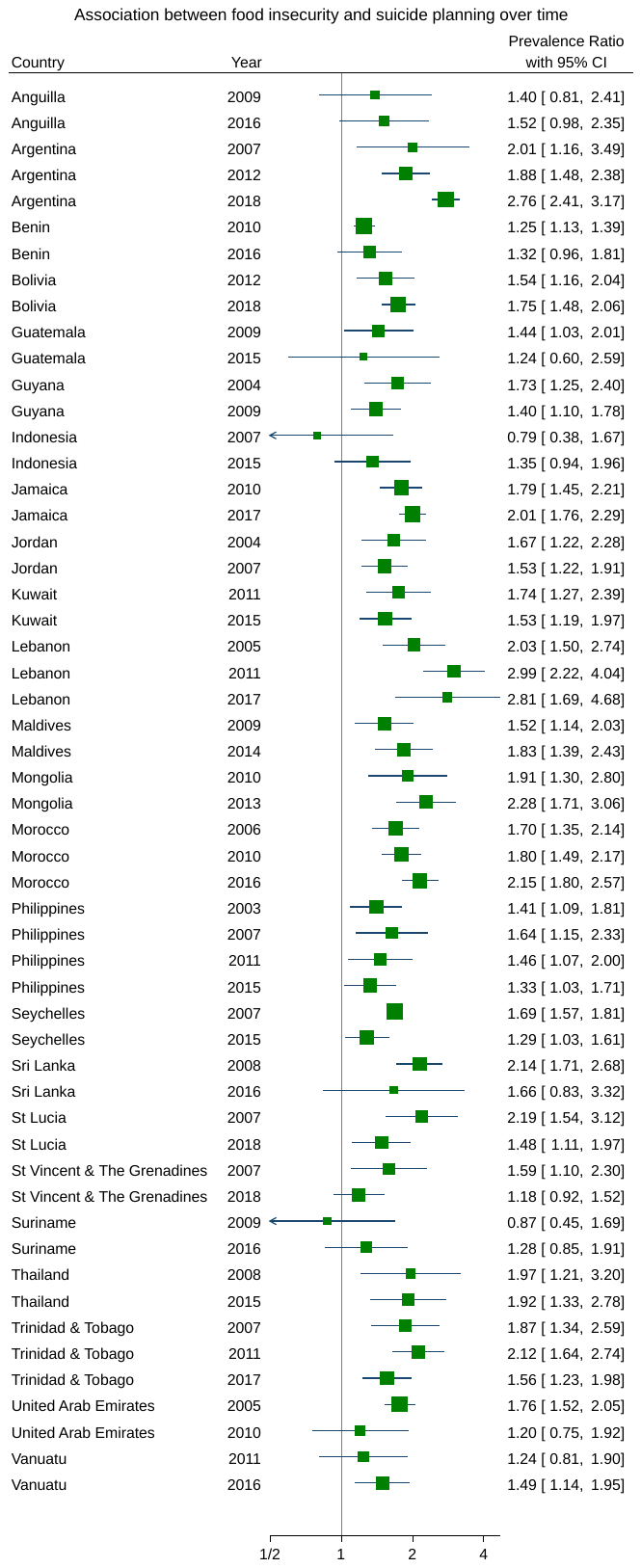

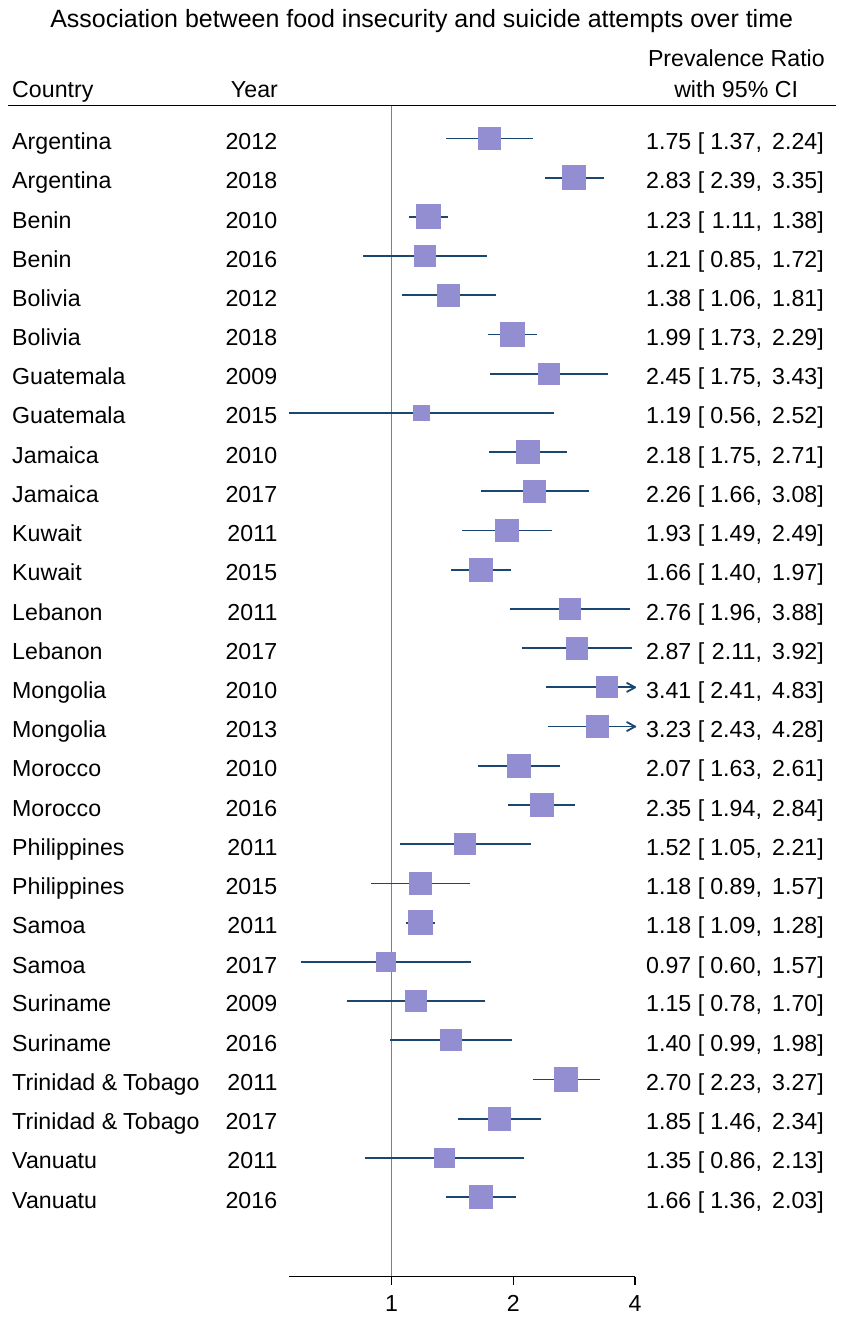
