## Supplementary material for "Food insecurity, adolescent suicidal thoughts and behaviours, and country-level context: a multi-country cross-sectional analysis": Figures

Figure 1. Forest Plots for the Relative Index of Inequality (RII) for suicidal ideation, suicide planning, and suicide attempts comparing the most food-secure adolescents with the least food-secure adolescents across countries and grouped according to WHO region.

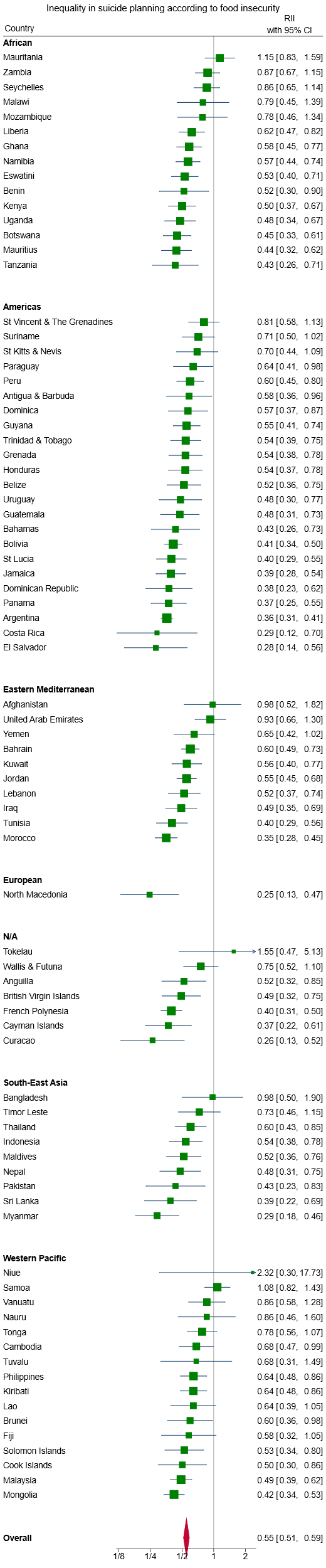

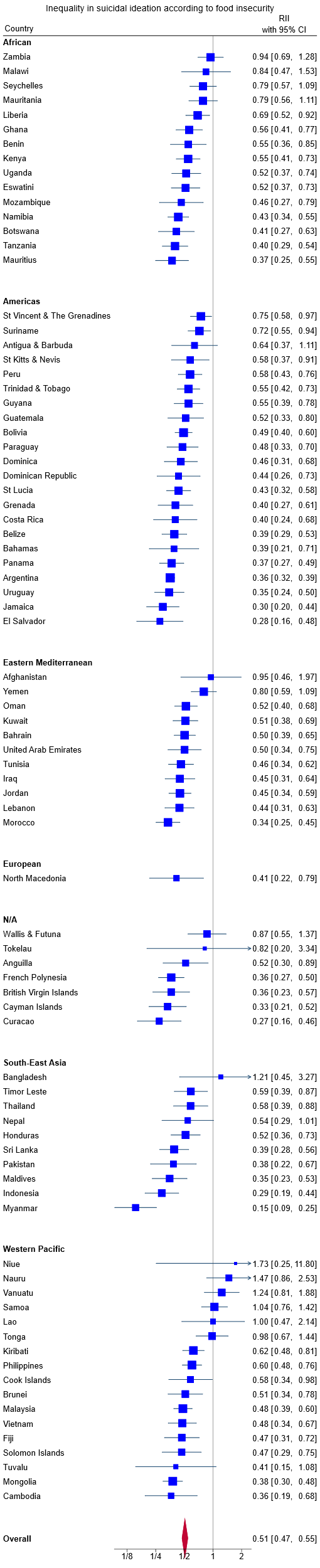

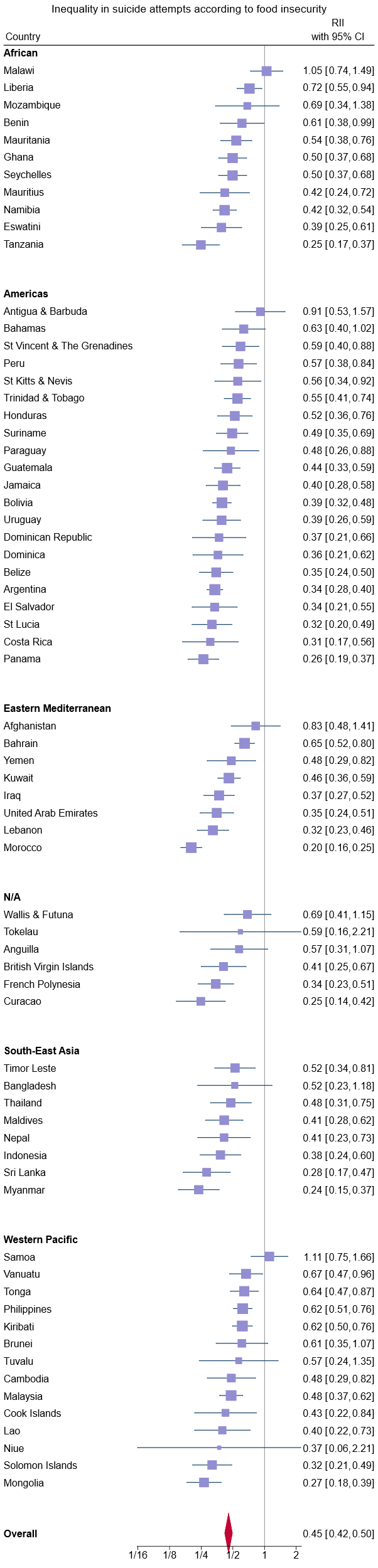

RIIs are plotted on a log axis. The vertical line at 1 on the x-axis represents no inequality in any outcome according to food insecurity status. Smaller numbers closer to zero indicate that the outcome is comparatively more prevalent in food-insecure adolescents. Larger numbers away from 1 indicate that the outcome is comparatively more prevalent in food-secure adolescents.

Figure 2. World maps for the Relative Index of Inequality (RII) for suicidal ideation, suicide planning, and suicide attempts comparing the most food-secure adolescents with the least food-secure adolescents across countries.

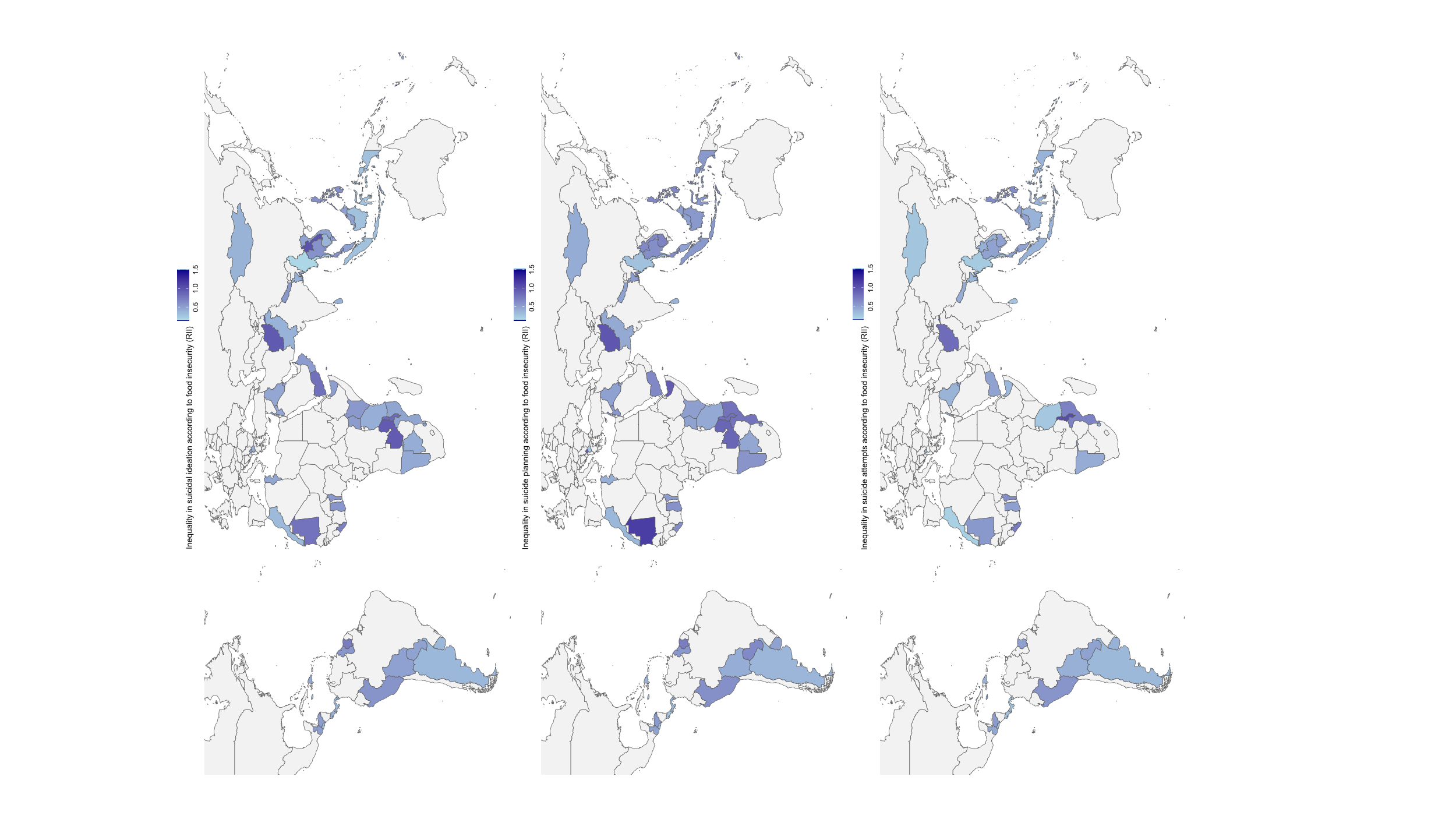

Lighter colours indicate that the outcome is comparatively more prevalent in food-insecure adolescents, with darker colours indicating smaller differences in the outcome according to food insecurity status or that the outcome is comparatively more prevalent in food-secure adolescents. Several small island countries and territories are not visible within the figure.

Figure 3. Bubble plots from the meta-regressions on the moderating effect of the national prevalence of adolescent food insecurity on the magnitude of the association between food insecurity (binary measure) and suicidal ideation, suicide planning and suicide attempt.

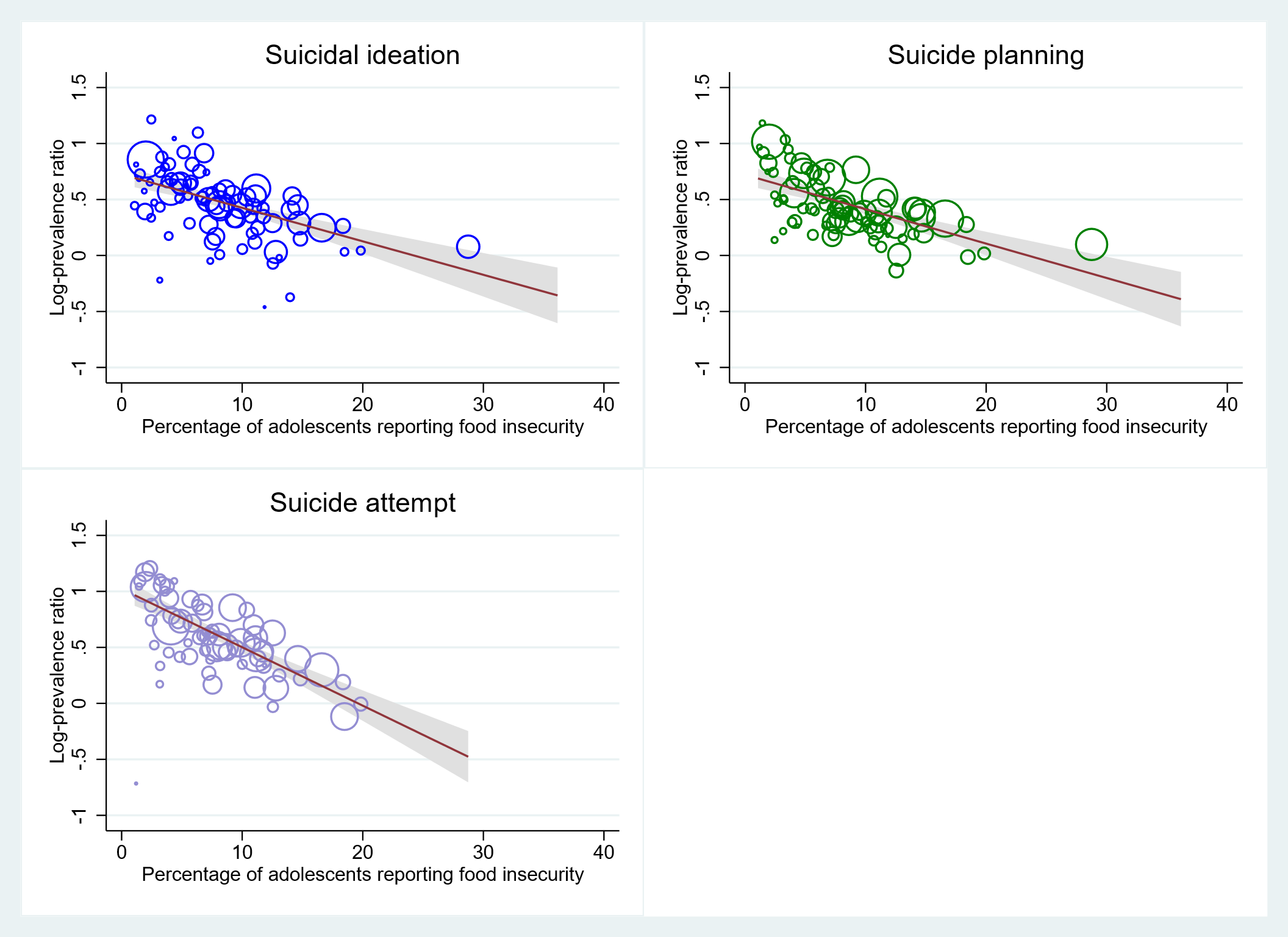

Each circle within the figure represents a country-level estimate with the size of the circle inversely proportional to the standard error. The solid line corresponds to the meta-regression estimate, and the corresponding 95% CI, indicated by grey shading.
